# Evaluating the impact of indoor residual spraying on malaria transmission in Madagascar using routine health data

**DOI:** 10.1101/2023.04.11.23288261

**Authors:** Emily R. Hilton, Saraha Rabeherisoa, Herizo Ramandimbiarijaona, Julie Rajaratnam, Allison Belemvire, Laurent Kapesa, Sarah Zohdy, Catherine Dentinger, Timothee Gandaho, Djenam Jacob, Sarah Burnett, Celestin Razafinjato

## Abstract

**Introduction:** Indoor residual spraying (IRS) and insecticide-treated bed-nets (ITNs) are cornerstone malaria prevention methods in Madagascar. This retrospective observational study uses routine data to evaluate the impacts of IRS overall, sustained IRS exposure over multiple years, and level of spray coverage (structures sprayed/found) in nine districts where non-pyrethroid IRS was deployed to complement standard pyrethroid ITNs from 2017 to 2020.

**Methods:** Multilevel negative-binomial generalized linear models were fit to estimate the effects of IRS exposure overall; consecutive years of IRS exposure; and spray coverage level on monthly all-ages population-adjusted malaria cases confirmed by rapid diagnostic test at the health facility level. The study period extended from July 2016 to June 2017. Facilities missing data and non-geolocated communes were excluded. Facilities in IRS districts were matched with control facilities by propensity score analysis. Models controlled for ITN survivorship, mass drug administration coverage, precipitation, enhanced vegetation index, seasonal effects, and district. Predicted cases under a counterfactual *no IRS* scenario and number of cases averted by IRS were estimated using the fitted models.

**Results:** Exposure to IRS overall reduced case incidence by an estimated 30.3% from 165.8 cases per 1,000 population (95%CI=139.7-196.7) under a counterfactual no IRS scenario, to 114.3 (95%CI=96.5-135.3), over 12 months post-IRS campaign in 9 districts. A third year of IRS reduced malaria cases 30.9% more than a first year (IRR=0.578, 95%CI=0.578-0.825, P<0.001) and 26.7% more than a second year (IRR=0.733, 95%CI=0.611-0.878, P=0.001). There was no significant difference between a first and second year (P>0.05). Coverage of 86%-90% was associated with a 19.7% reduction in incidence (IRR=0.803, 95%CI=0.690-0.934, P=0.005) compared to coverage ≤85%, although these results were not robust to sensitivity analysis.

**Conclusion:** This study demonstrates that non-pyrethroid IRS appears to substantially reduce malaria incidence in Madagascar and that sustained implementation of IRS over 3 years confers additional benefits.

**KEY MESSAGES:** *What is already known on this topic:* The use of non-pyrethroid insecticides for indoor residual spraying (IRS) in communities using pyrethroid-based insecticide treated nets (ITNs) is associated with reduced malaria prevalence, and with reduced malaria incidence in some settings.[1] To date, few studies have investigated the impact of high spray coverage and sustained IRS implementation over multiple years.[2,3]

*What this study adds:* This study estimates the impact of IRS in a setting with heterogenous malaria transmission and variation in intervention impact at the subnational level. Additionally, this study presents evidence of a benefit to continuing IRS implementation over multiple consecutive years.

*How this study might affect research, practice, or policy:* The results reported here bolster the evidence around the effectiveness of non-pyrethroid insecticides for IRS while suggesting that policymakers should consider the benefits of sustaining IRS implementation over multiple years when undertaking decisions to move locations and/or withdraw IRS.

## INTRODUCTION

Malaria prevalence worldwide has declined in recent decades, with significant contributions from the implementation of vector control interventions including indoor residual spraying (IRS).[4] However, in recent years progress has stalled and in some areas reversed.[5] Malaria is among the top five causes of mortality in Madagascar,[6] and in 2020 the country accounted for 1.5% of global malaria cases and 1.5% of global malaria deaths. From 2015 to 2020, Madagascar experienced an increase in malaria incidence and mortality greater than observed in other countries of East and Southern Africa.[5]

IRS is an intervention that involves applying internal walls of homes with a residual insecticide that will kill mosquitoes and other insects that encounter the treated walls. From 1993 to 1998, widespread IRS campaigns using DDT carried out in the Central Highlands of Madagascar successfully reduced malaria prevalence and vector abundance in the sprayed areas.[7] From 1999 to 2005, targeted DDT-based IRS operations continued in the Central Highlands. In 2005, the Malagasy National Malaria Control Programme (NMCP) replaced DDT with pyrethroid insecticides for IRS in the Highlands.[8] Since 2008, the U.S. President’s Malaria Initiative (PMI) has supported IRS campaigns in Madagascar, most recently through the PMI VectorLink Project (2017–2023) and its predecessor, the Africa Indoor Residual Spraying (AIRS) program (2014–2017). From 2008 to 2012, “generalized” IRS spraying, meaning close to 100% of eligible structures are sprayed, was deployed in the pre-elimination Central Highlands area. In 2012, the strategy was shifted to “focalized” spraying, which targeted areas in the Central Highlands with higher malaria case incidence, an approach that was sustained through 2015. Starting in 2014, the country shifted to a strategy of targeting high-transmission regions and deployed IRS in the east and southeast coastal districts.[9] IRS has since been targeted to select high-burden areas based on epidemiological and entomological data, as well as other contextual factors such as the presence of environmentally protected areas and organic agriculture.[10] Malaria vector resistance to pyrethroids has been detected in eastern and southeastern coastal regions,[11] and from 2016 to 2020 non-pyrethroid insecticide products including pirimiphos-methyl and clothianidin were deployed. The 2018– 2022 National Strategic Plan in Madagascar recommends rotation of insecticides every 2 to 3 years to manage insecticide resistance.[6]

Although IRS is a widely used core vector control intervention that has been implemented to successfully reduce malaria incidence[12–15] for decades, increasing resistance to pyrethroids presents a major challenge in global progress against malaria.[16] The World Health Organization (WHO) Global Plan for Insecticide Resistance Management has encouraged the use of non-pyrethroid insecticides for IRS to reduce the spread of insecticide resistance,[17] although rigorous evidence on the effectiveness of next-generation non-pyrethroid IRS products on real-world clinical outcomes is lacking.

WHO recommends that IRS structure coverage of at least 85% be achieved in order to attain a mass effect on local vector populations.[18] Although there is a strong theoretical foundation for ensuring high coverage of IRS, to date few reports have shown evidence for the relationship between level of coverage at the community level and reduction in malaria burden.

Given the cost of IRS implementation, NMCPs and donors often must make crucial targeting decisions for IRS campaigns, including whether to sustain implementation in the same place over several years or to rotate targeted geographies. This decision rests on two questions: (1) what is the impact of withdrawing IRS after one or multiple rounds of implementation, and (2) is there additional benefit to implementing IRS over multiple consecutive years in the same area? The impact of withdrawing IRS has been previously investigated.[3,19]

Here we present an observational, retrospective analysis of the epidemiological impact of IRS in 9 total districts of Madagascar where IRS was implemented from 2017 to 2020. This study has three objectives: (1) to estimate the overall impact of IRS campaigns on routinely reported malaria incidence; (2) to investigate the effect of implementing IRS over multiple consecutive years; and (3) to estimate the impact of high IRS coverage that meets the WHO 85% target. An additional aim of this work is to foster the use of routine malaria surveillance data for evidence-based decision-making.

## METHODS

### Study setting

Madagascar is a tropical island located in the southwest of the Indian Ocean, approximately 400 km off the western coast of Mozambique. The country is ecologically and climatically diverse, reflected in the highly heterogenous nature of malaria transmission and seasonal malaria patterns across the island.[20] Malaria has increased in all areas of Madagascar since 2011,[21] with high endemicity observed chiefly in the eastern and western coastal regions, and lower transmission in the Central Highlands.[22] IRS deployment from 2017 to 2020 took place in southeastern and southwestern districts (Figure 1) in regions with primarily tropical climates, except for two districts (Betioky Atsimo and Ampanihy Ouest) located in a region where the climate is classed as subdesert. Malaria prevalence in children aged 6-59 months measured in the 2016 Malaria Indicator Survey (MIS) was found to be 14.9% in the tropical areas and 5% in the subdesert areas.[23] The 2021 Demographic Health Survey (DHS) found that prevalence had increased to a range of 15.8% to 27.2% prevalence in tropical regions where IRS was deployed, and decreased to 4.2% in the subdesert.[24] The DHS survey reported that 69% of households nationally had access to at least one insecticide-treated bed net (ITN), a decrease from 80% reported by the 2016 MIS.[23,24] Administratively, the country is divided into 23 regions and 114 districts, which are further subdivided into communes.

**Figure 1.**
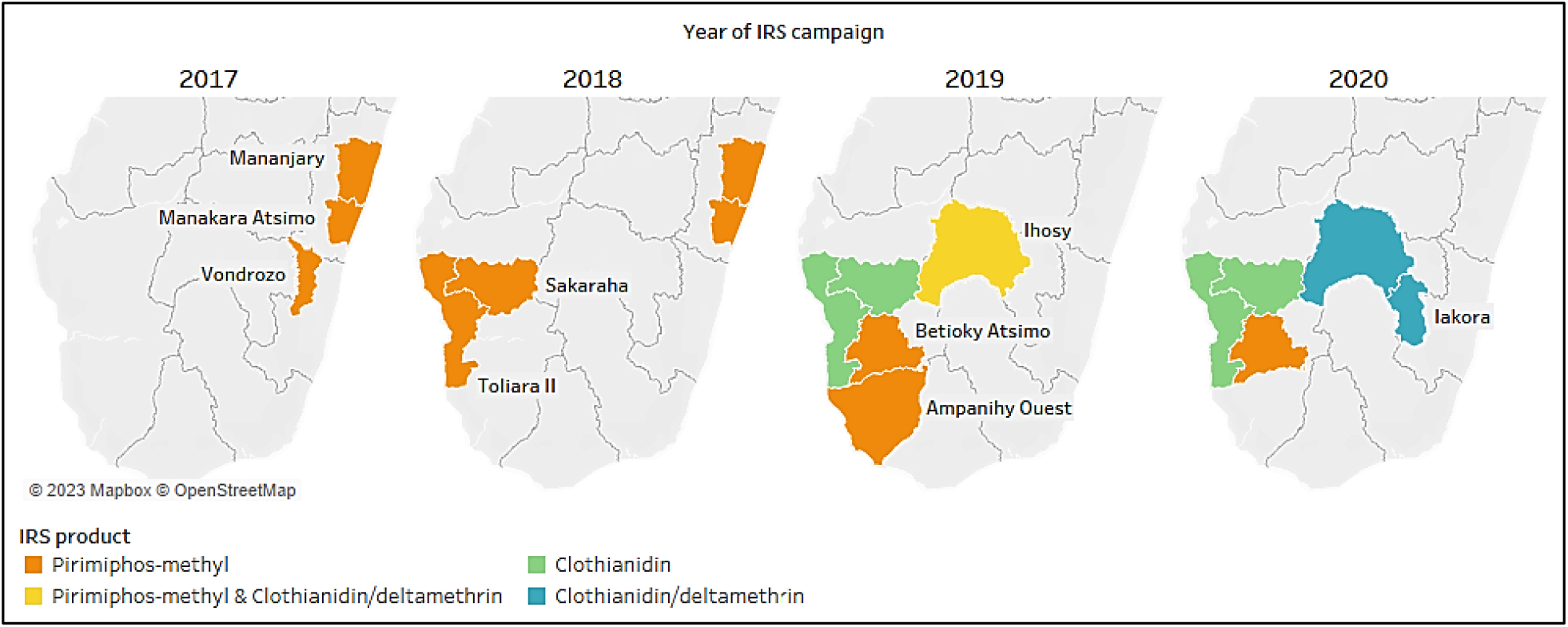
Map of districts where indoor residual spraying (IRS) campaigns were implemented from 2017 to 2020 and the IRS products used.

### Intervention

From 2014 through 2020, PMI VectorLink/AIRS deployed annual IRS spray campaigns in 14 total districts of Madagascar. Due to concerns around HMIS data quality prior to 2016, 5 districts where IRS was deployed from 2014 to 2016 were excluded from this study, leaving 9 study districts. Between 3 and 5 study districts were targeted for IRS each year. Each study district received between one and three consecutive years of IRS implementation with pirimiphos-methyl (Actellic® 300CS), clothianidin (SumiShield® 50WG), or a combination of clothianidin and deltamethrin (Fludora® Fusion) (Figure 1). IRS coverage was reported at the commune level (the administrative level below district). IRS campaigns took place from July to September in 2017, and 2018; and from October to December in 2019 and 2020. Of the 9 study districts that received at least one year of IRS implementation from 2017 to 2020, 6 received a second consecutive year, and 3 received a third year.[25–29]

### Study design

This study is a retrospective observational evaluation of the epidemiological impact of IRS campaigns that took place from 2017 to 2020 in 9 districts of Madagascar. The evaluation period encompassed July 2016 through June 2021.The primary outcome was confirmed outpatient malaria cases among all ages, standardized per health facility catchment population as reported to the health management information system (HMIS). Case confirmation was done by rapid diagnostic testing (RDT). Exposure was defined at the commune level (administrative level below district) by IRS status (whether IRS was conducted within the commune) and by level of spray coverage achieved. Further details on IRS exposure definitions are provided in the following sections. The evaluation used a “transmission year” defined as starting in July and extending to June of the following year to account for seasonal malaria transmission.

### Primary outcome

The primary outcome was monthly RDT-confirmed outpatient malaria cases among all ages, standardized per health facility catchment population. Malaria case data were extracted from the Madagascar HMIS for all health facilities. In 2017 Madagascar’s HMIS transitioned to the DHIS2 platform from a previous database based in Microsoft Access. Data from January 2016 onward were available in DHIS2. The final dataset used in this study was accessed from DHIS2 on October 12, 2021, for the period July 2016 to June 2021, including malaria cases confirmed by RDT and health facility catchment populations. Microscopy testing and unconfirmed cases are not reported in DHIS2. Facility catchment populations are defined based on the most recent census conducted in 2018 and projected forward and backward assuming a 2.9% annual growth rate.[30]

Health facilities were excluded from analysis according to the following criteria: private health facilities and hospitals; facilities missing catchment population data; facilities in communes missing geocoordinates, which did not allow them to be linked to climate data; and facilities missing RDT-confirmed case data for an entire year. Facilities in five districts that had received IRS from 2014 to 2016 were excluded. Anomalous values were identified by visual inspection of time-series plots for each health facility and comparing against outpatient totals. One value determined to be anomalous for being greater than four standard deviations from the mean was excluded.

### Potential confounding variables

To account for the potential impact of other malaria control interventions on the outcome, indicators of access to ITNs and mass drug administration (MDA) coverage were included in the analysis.

Mass distribution of standard pyrethroid ITNs took place in October and November 2015 in 92 districts and in August 2018 in 106 districts. Both distributions included all 9 IRS study districts. ITN survivorship was included as a covariate to control for the time since the most recent ITN campaign, based on durability monitoring conducted by the PMI VectorLink Project and the Institut Pasteur de Madagascar.[31,32] Durability monitoring using standard protocols [33,34] found that net survivorship declined exponentially after both distributions during the 36-month monitoring period. Net survivorship was averaged at each follow-up time point (3–6 months, 12 months, and 24 months post-distribution in 2015; 12 months, 24 months, and 36 months post-distribution in 2018) and applied monthly to districts based on the time since the previous ITN campaign. Net survivorship was set to zero for districts that were not included in the ITN campaigns.

Two rounds of MDA with dihydroartemisinin-piperaquine took place in 11 districts in March and June 2021. Of the 11 districts where MDA took place, only one district, Ampanihy Ouest, had also received IRS implementation (in 2019). MDA coverage data at the district level was provided by the Madagascar NMCP. Coverage was calculated as the number of treated inhabitants divided by total population and included in analysis as a continuous covariate.

Monthly precipitation data were pulled from the Climate Hazards Group InfraRed Precipitation with Station (CHIRPS) dataset[35] and enhanced vegetation index (EVI) data were pulled from the Famine Early Warning Systems Network (FEWS NET).[36] Commune-level shapefiles were used to access monthly averages from these spatial datasets, which were matched by name to commune-level data from the HMIS. To account for varying climate and transmission patterns across Madagascar, precipitation and EVI lags were calculated using Pearson correlation tests with malaria cases, with districts grouped into the eight malaria transmission ecozones identified by Howes et al.[37] The selected lagged values were then scaled to have a mean of zero and a standard deviation of one.

### Analytical approach

#### Matching intervention and control groups

Selection of control groups was done using propensity score matching, a method for obtaining intervention and control groups with similar covariate distributions.[38] Matching was performed at the health facility level, and all health facilities in non-IRS districts meeting the inclusion criteria described above were eligible to be matched as controls. For each facility, EVI, precipitation, ITN survival, and RDT-confirmed malaria case incidence per 1,000 population were averaged over a 12-month baseline period (July 2016 to June 2017) before IRS had been deployed in any of the study IRS districts. Propensity scores were obtained via logistic regression where the previously described variables were included as covariates. The outcome was a binary variable for which facilities that would receive IRS were assigned a one and facilities that would not were assigned a zero.

IRS and control facilities were matched based on their propensity scores using nearest neighbor matching with a one to four ratio (each IRS facility was matched to four control facilities) and without replacement (each control facility could only be matched with one IRS facility). Diagnosis of the quality of matches was done by comparing covariate distributions in control and IRS groups. The standard mean difference (SMD) for each covariate was assessed against a threshold of 0.25, and density plots and distribution plots of propensity scores were examined.[39]

#### Model development

Different models were developed to respond to each study question and estimate the effect of IRS exposure on the outcome variable. The models differed primarily according to their exposure variables as described in the sections below. For each study question a negative binomial generalized linear multilevel mixed effects (GLMM) model with a log link was specified, with the following basic formulation:

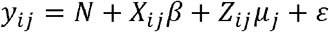

Where *y*_*ij*_ is malaria cases at the *i*^*th*^ month in the *j*^*th*^ cluster, *X* is the *p* × 1 row vector of *p* fixed-effects predictor covariates, and *β* is a *p* × 1 column vector of the fixed-effects regression coefficients. The *q* × 1 vector of *q* random-effects covariates is given by *Z*_*ij*_, and their coefficients by *μ*_*j*_, where the *j* subscript indexes the random effects grouping variables. The vector of residuals is given by *ϵ*; and an offset of the natural log of facility population is given by *N*. Variables included as fixed and random effects in each model are described in the following sections. Coefficients for the exposure variables of each model were exponentiated to obtain the incidence rate ratio (IRR).

#### Study question 1: Overall impact of IRS

To estimate the overall impact of IRS exposure on all-ages RDT-confirmed malaria incidence, the model dataset included all health facilities in IRS districts and their comparator facilities selected through propensity score matching.

IRS exposure was indicated using two binary dummy variables applied at the commune-month level. These variables indicated the first 6 months and the 7th to 12th months since the most recent IRS campaign. For the “IRS exposure 0-6 months” variable, commune-months within the first six months post-IRS were assigned a one and all others were assigned a zero. Similarly, for the “IRS exposure 7-12 months” variable, commune-months within seven to 12 months post-IRS were assigned a one and all others were assigned a zero. The 0–6 and 7–12 -month split of the variables reflects the reported residual efficacy of IRS, which tends to wane after 6 months.[40] In the districts Sakaraha and Toliara II, where 13 months passed between the 2018 and 2019 IRS campaigns, the 13th month was also assigned a “1” for the 7–12-month IRS status variable, to reflect the sustained IRS implementation in those districts.

IRS exposure variables along with ITN survival, MDA coverage, precipitation, EVI, and district were included in the model as fixed effects. Seasonal effects of malaria were accounted for by including cosine and sine functions, which assume that seasonal patterns follow a cosine function with variable amplitude and horizontal shift, and can be described as follows:

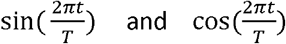

where *t* is the month and *T* is 12 (number of months in a year).[41] District and transmission year were interacted with the sine term to accommodate for variations in seasonality by district and over the course of the study period. Transmission year and clusters of matched IRS facilities with their comparator facilities were included as random effects. Interactions between district and other covariates were tested and the final model was selected based on having the lowest Akaike information criterion (AIC) score.

To quantify the model results in a more meaningful measure of overall impact of IRS, the number of malaria cases averted by IRS was estimated. The final fitted model was used to predict malaria cases under a counterfactual scenario of *no IRS* and compared against predicted cases under observed conditions. This step also allowed the inference of findings for areas missing RDT-confirmed case data and to generate district-specific results. The counterfactual prediction dataset was created by duplicating the model-fitting dataset and setting all IRS exposure variables to zero. The variables in the prediction dataset under observed conditions were unchanged. Both prediction datasets included facility-months missing RDT-confirmed case data, which had previously been excluded from model-fitting. Monthly malaria case counts in each of the nine IRS districts were estimated using the fitted model and the prediction datasets. Uncertainty intervals were generated by simulating from the fixed (betas and associated variance-covariance matrix) and random (predicted random effect and associated standard error) parts of the model to generate 1,000 linear predictions per facility-month for both the observed and counterfactual scenarios. Cases averted by IRS were calculated by subtracting predicted cases under observed conditions from predicted cases under the *no IRS* counterfactual scenario. The mean of these 1,000 simulations was the resulting point estimate, and the 2.5th and 97.5th percentiles were the lower and upper bounds of the uncertainty intervals.[42]

#### Study question 2: Impact of multiple years of exposure to IRS

A model structure similar to that used in study question 1 was used to estimate the impact of sustained exposure to IRS over multiple years, with differences in the IRS exposure variables and seasonal effects. Three binary IRS exposure variables were applied to the commune-spray-year. A spray-year was defined as the 12 months following the start of IRS implementation. These variables indicated the 12 months following an IRS campaign and whether that campaign was a first, second, or third consecutive year of IRS implementation. In districts where 13 months passed between spray campaigns, the 13th month was considered part of the preceding spray year. Month and transmission year were included in the model as fixed effects in the place of the sine and cosine terms used above, as a result of improved model fit determined by low AIC score. Estimates for linear combinations of the three IRS exposure variables were computed to determine if there was a significant difference between estimates.

#### Study question 3: Impact of level of IRS spray coverage

To estimate the impact of level of spray coverage, the modeling dataset included only data from the nine IRS districts, during the 12 months following an IRS campaign. Two models were developed with structures similar to those described above and with differences as follows: IRS exposure, defined as spray coverage (the number of structures sprayed out of structures found) was applied to the level of commune-spray-year. Spray coverage was first modeled as a categorical dummy variable with four coverage bins: <85%, 85%–90%, 91%–95%, and 96%–100%, to determine if significant benefit is detected at the WHO-recommended 85% threshold or at a higher level of coverage. Second, a sensitivity analysis was conducted with spray coverage modeled as a continuous covariate to examine a potential linear relationship with the outcome variable. These models did not include a covariate for MDA coverage, as this intervention did not take place in any of the IRS districts in the same year as an IRS campaign. Clusters of matched IRS and comparator facilities were not included as random effects, as the modeling datasets did not include any comparator data. Communes were instead included as random effects.

Data were cleaned, transformed, and joined in R version 4.1.1 and Alteryx Designer x64 version 2021.1 (Alteryx, Inc., Colorado, USA). Model fitting and simulations were run in Stata/SE 17.0 (StataCorp LLC, Texas, USA). Data visualizations were created in Tableau version 2021.2.4 (Tableau Software LLC, Washington, USA).

### Data sources

Routinely collected data on IRS implementation and coverage were sourced from the PMI AIRS and the PMI VectorLink projects. Malaria case data and health facility populations were extracted from the Madagascar HMIS. ITN and MDA campaign data and administrative shapefiles were provided by the Madagascar NMCP.

### Patient and Public Involvement

Patients and the public were not involved in this research study.

## RESULTS

A total of 2,547 public health facilities were identified in the Madagascar HMIS during the study period (July 2016 to June 2021), of which 2,171 met the eligibility criteria for inclusion in this study. Of these, 214 facilities (9.8%) were in IRS districts. After matching based on propensity scores, 848 control facilities from 76 districts were retained for inclusion in modeling for study questions one and two. Only the 214 facilities in IRS districts were included in modeling for study question three. (Further details on facility inclusion in the analysis dataset can be found in the supplemental material.)

Epidemiological curves showing the average monthly incidence of RDT-confirmed malaria cases of facilities in IRS districts and their matched comparator facilities are shown in Figure 2**Error! Reference source not found**.. In the final analysis dataset, a total of 3,412,689 RDT-confirmed malaria cases were reported during the study period, of which 775,547 (22.7%) were reported from facilities in IRS districts. A wide range of case incidence was observed throughout the study period. Among IRS districts, district-level annual incidence ranged from 8.6 to 435.1 confirmed cases per 1,000 population.

**Figure 2.**
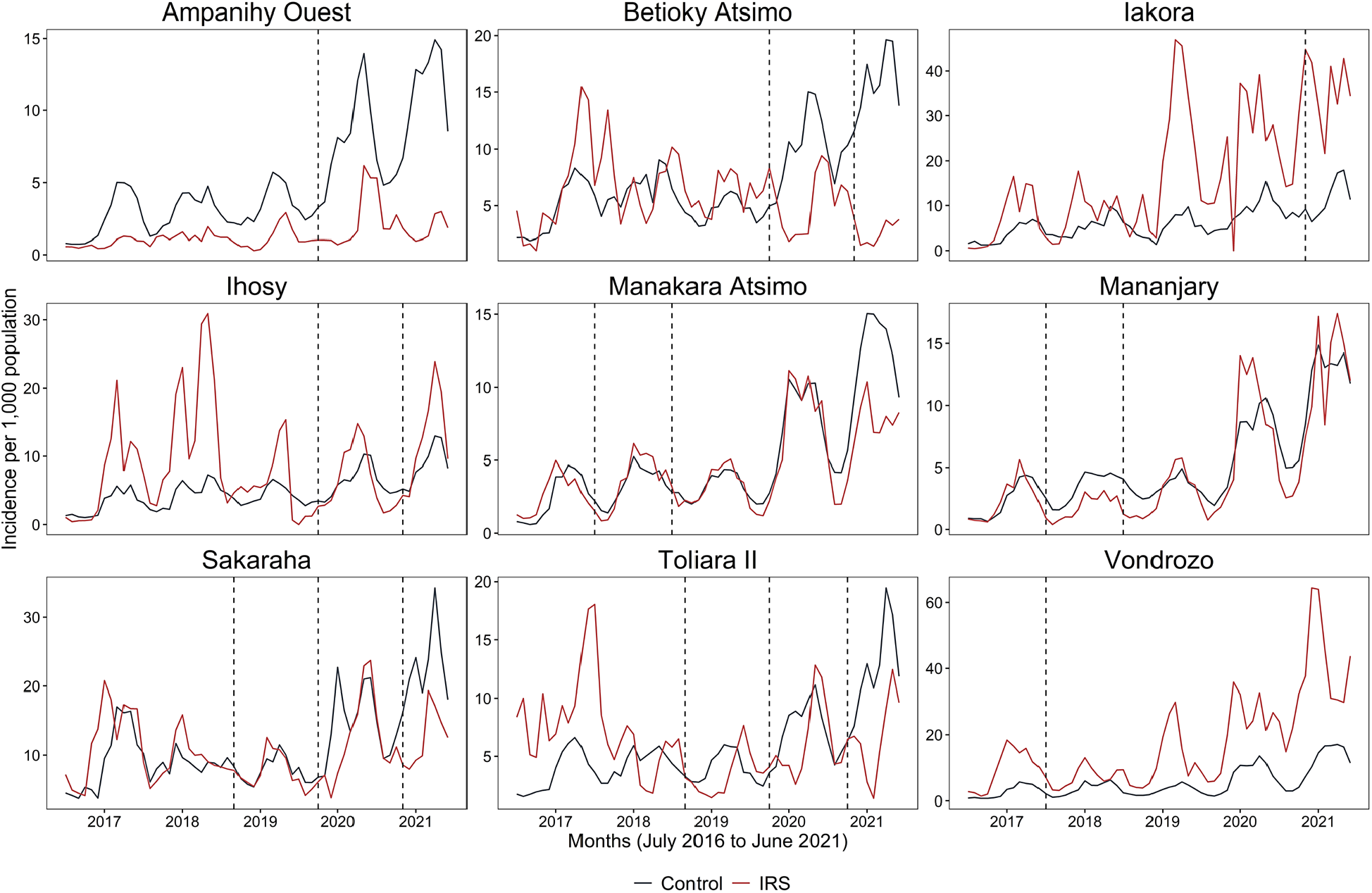
Monthly incidence per 1,000 population of routinely reported malaria cases (confirmed by rapid diagnostic test) in the 9 indoor residual spraying (IRS) study districts from July 2016 to June 2021 and their matched controls. The dates of IRS campaigns are indicated by vertical dashed lines. Red lines represent facilities in IRS districts and black lines represent matched control facilities.

Summary characteristics and IRS campaign results as well as the final lags for rainfall and EVI included in modeling for each malaria transmission ecozone are available in the supplementary material.

### Study Question 1: Overall impact of IRS

The first 6 months following an IRS campaign were associated with an estimated 43.3% reduction in malaria incidence (IRR=0.567, 95%CI=0.531-0.605; P<0.001); and the 7th to 12th months post-IRS showed a 24.8% reduction in incidence (IRR=0.752, 95%CI=0.704-0.802; P<0.001) (Figure 3).

**Figure 3.**
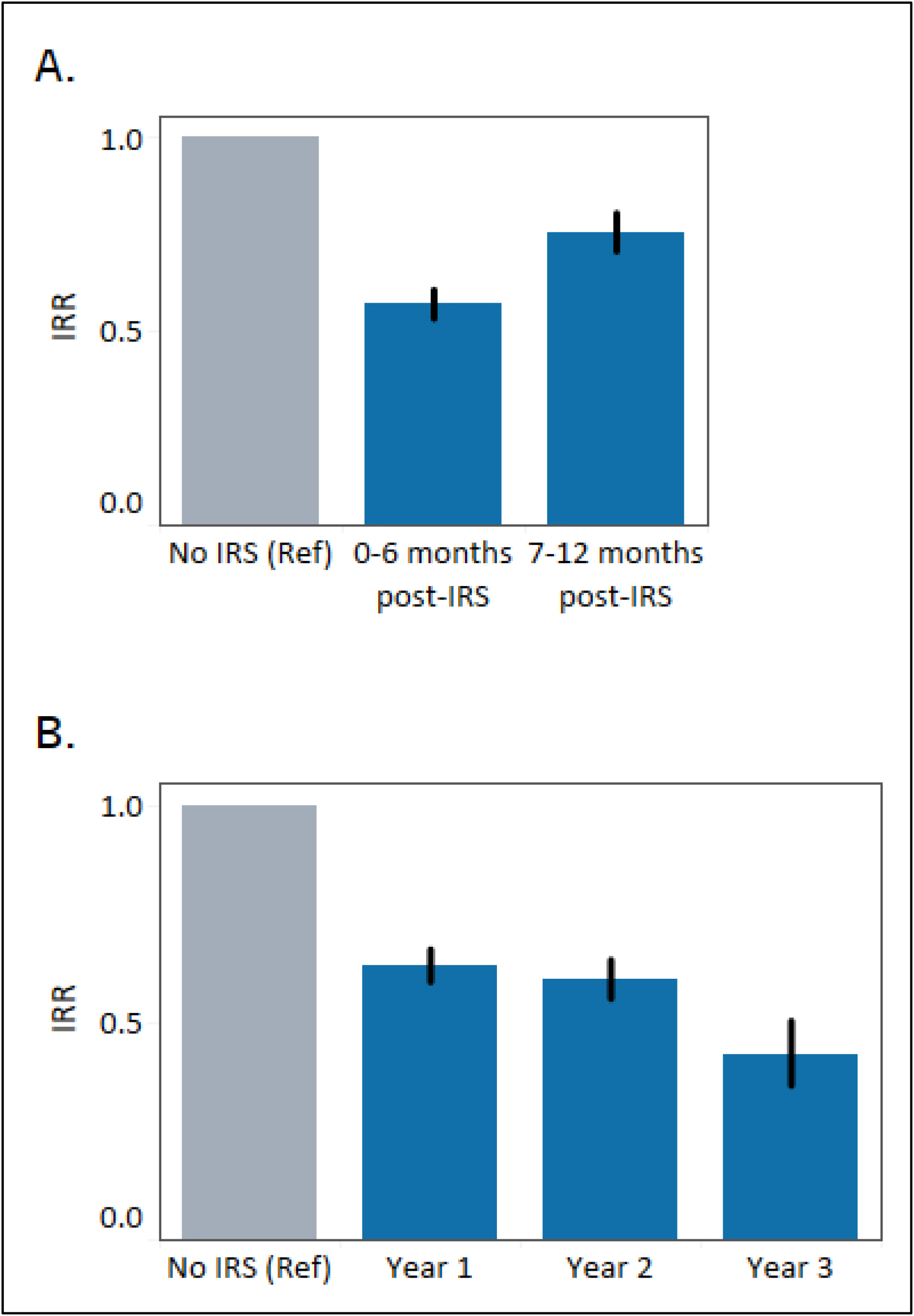
Incidence rate ratio (IRR) results from two regression models describing the association between number of RDT-confirmed malaria cases among all ages and (A) indoor residual spraying (IRS) status 0–6 or 7–12 months following an IRS campaign; (B) IRS exposure in the 12 months following the first, second, or third consecutive year of IRS implementation. Asterisks indicate P<0.05.

An estimated 116,323 (95%CI=40,152-196,764) cases of malaria were averted between July 2016 and June 2021 by IRS campaigns in the 9 study districts. Malaria incidence was reduced by an estimated 30.3% from 165.8 (95%CI=139.7-196.7) cases per 1,000 population under a counterfactual no IRS scenario, to 114.3 (95%CI=96.5-135.3) under observed conditions, over the 12 months following an IRS campaign in the 9 spray districts. The estimated incidence of cases averted per 1,000 population was highest in Sakaraha following the 2020 campaign (56.52, 95%CI=11.29-105.72) and lowest in Mananjary following the 2017 IRS campaign (5.26, 95%CI=0.52-10.07). Estimated incidence of cases averted was not significant in Iakora (245.88, 95%CI=-46.86-567.69) following the 2020 IRS campaign. A map of estimated cases averted per 1,000 population for each district where IRS campaigns were implemented is shown in Figure 4 along with graphs of modeled case incidence under observed and counterfactual scenarios. The full model regression results are available in the supplementary material.

**Figure 4.**
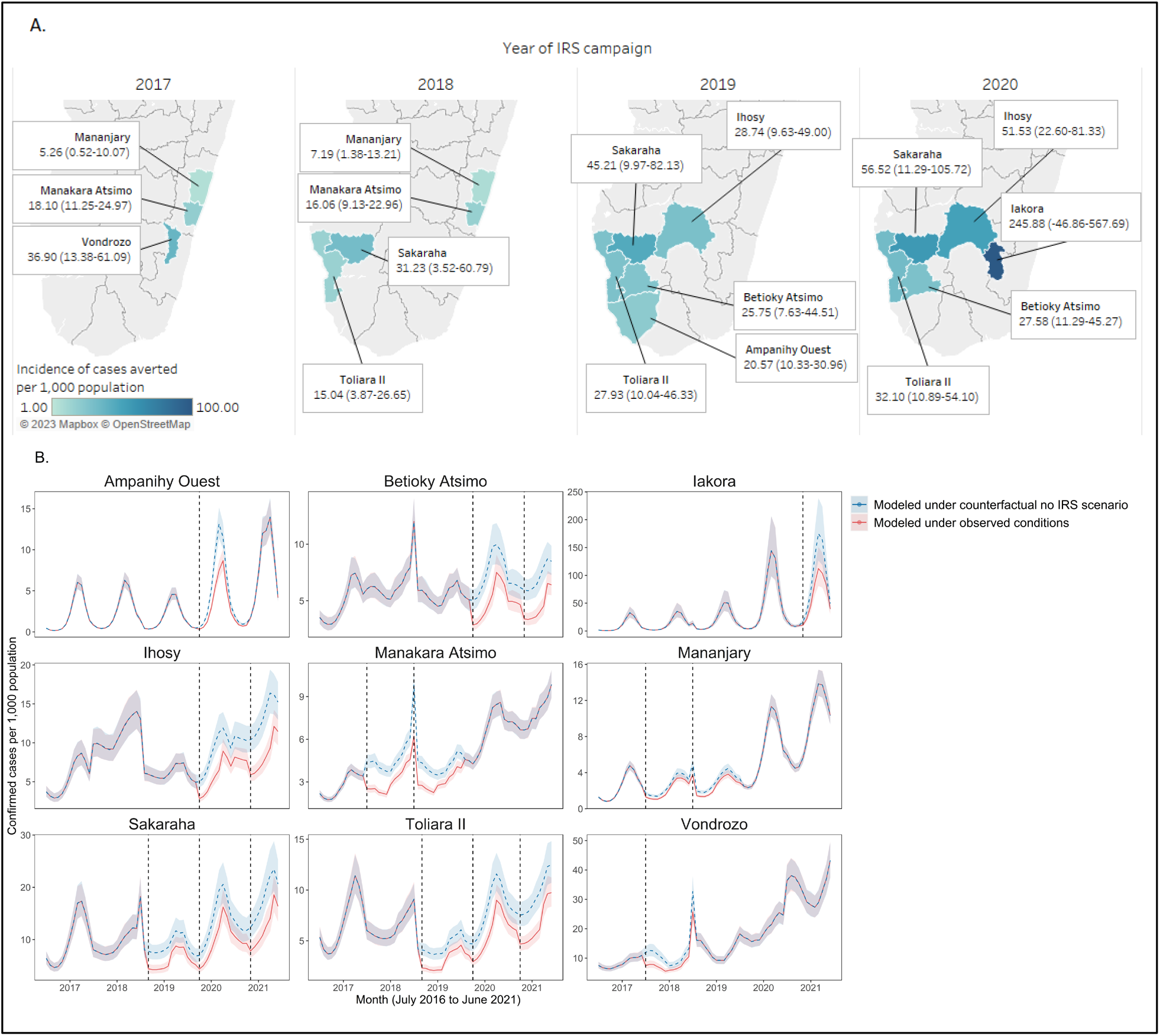
Estimated malaria cases averted per 1,000 population with 95% confidence intervals in nine districts where indoor residual spraying (IRS) campaigns were implemented. (A) Map of incidence of cases averted shown by year of spray campaign. Cases averted are estimated over the 12 months following IRS implementation. Map labels show point estimates with 95% confidence intervals in parentheses. (B) Estimated monthly case incidence in IRS districts from July 2016 to June 2021 under observed conditions (red) and under a counterfactual “no IRS” scenario (blue). Shaded areas around the solid lines represent 95% confidence intervals. IRS campaign dates are indicated by vertical dashed lines.

### Study Question 2: Impact of multiple years of exposure to IRS

A single year of IRS exposure was associated with a 37.0% reduction in malaria case incidence (IRR=0.630, 95%CI=0.594–0.668, P<0.001), and a second year of IRS was associated with a 40.2% reduction in incidence (IRR=0.598, 95%CI=0.555–0.644, P<0.001) compared with no IRS. Compared with the first year of IRS, a second year was not associated with a significantly different reduction in incidence (IRR=0.943, 95%CI=0.866-1.023, P=0.170). IRS in year 3 continued to reduce case rates compared to no spray (IRR=0.423, 95%CI=0.355–0.504, P<0.001), and produced a significantly greater effect than both year 1 and year 2. The year 3 percent reduction in cases was 30.9% greater than the effect in year 1 (IRR=0.578, 95%CI=0.578-0.825, P<0.001) and 26.7% greater than in Year 2 (IRR=0.733, 95%CI=0.611-0.878, P=0.001) (Figure 3). The full model regression results are available in Supplement 4.

### Study Question 3: Impact of level of IRS spray coverage

In 2019 and 2020, all communes achieved spray coverage at or above 85%. From 2017 to 2018, 22 of 300 total communes (7.3%) achieved coverage at or below the 85% target threshold set by WHO**Error! Reference source not found**.. Low coverage was generally attributable to accessibility challenges in remote areas and refusals.[25–29] Twenty-two communes (7.3%) achieved coverage between 86% to 90%; 86 communes (28.7%) achieved coverage from 91% to 95%; and 170 communes (56.7%) achieved coverage higher than 95% (Table 1).

**Table 1.**
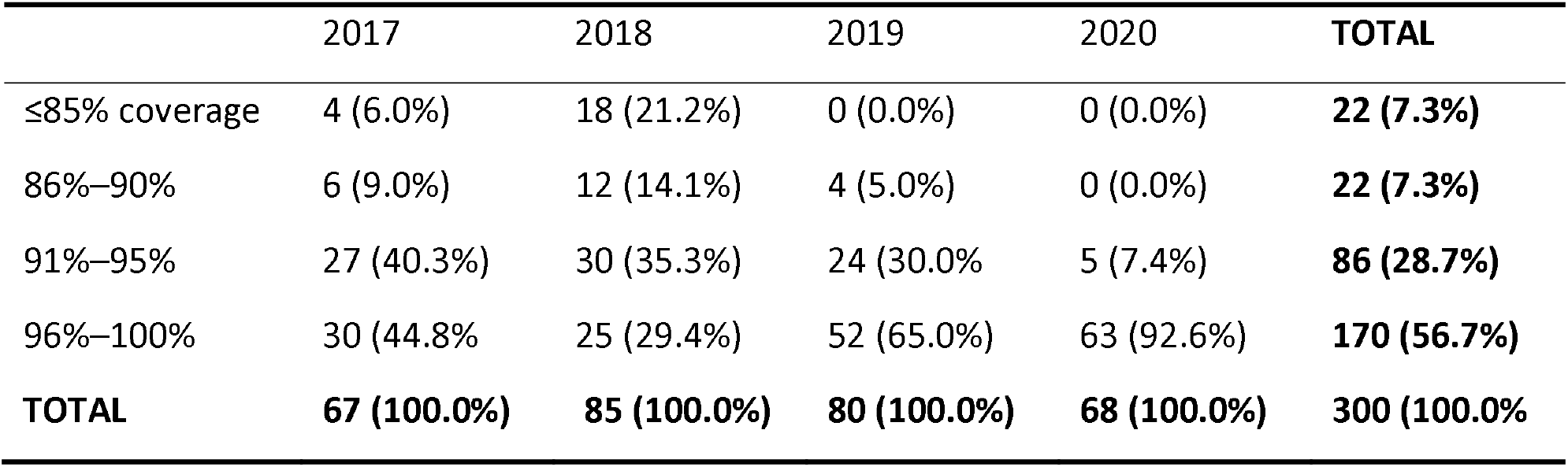
Number and percentage of communes that achieved each indoor residual spraying (IRS) coverage threshold in campaigns that took place from 2016 to 2020.

|  | 2017 | 2018 | 2019 | 2020 | TOTAL |
| --- | --- | --- | --- | --- | --- |
| ≤85% coverage | 4 (6.0%) | 18 (21.2%) | 0 (0.0%) | 0 (0.0%) | **22 (7.3%)** |
| 86%–90% | 6 (9.0%) | 12 (14.1%) | 4 (5.0%) | 0 (0.0%) | **22 (7.3%)** |
| 91%–95% | 27 (40.3%) | 30 (35.3%) | 24 (30.0%) | 5 (7.4%) | **86 (28.7%)** |
| 96%–100% | 30 (44.8%) | 25 (29.4%) | 52 (65.0%) | 63 (92.6%) | **170 (56.7%)** |
| **TOTAL** | **67 (100.0%)** | **85 (100.0%)** | **80 (100.0%)** | **68 (100.0%)** | **300 (100.0%)** |

When modeled as a categorical variable, IRS spray coverage between 86% and 90% was associated with a 19.7% reduction in incidence (IRS=0.803, 95%CI=0.690-0.934, P=0.005) compared to spray coverage at or below 85%. IRS spray coverage in categories higher than 90% were associated with smaller and non-significant (p>0.05) reductions in incidence compared to coverage at or below 85%. When modeled as a continuous covariate, IRS spray coverage was significantly associated with a 1.0% increase in malaria incidence with every one percent increase in coverage (IRR=1.010, 95%CI=1.003-1.016, P=0.003). Full model regression results are available in the supplemental material.

## DISCUSSION

This study presents evidence of the impact of IRS on reduction of malaria burden in Madagascar. Our results demonstrate a consistent overall association between implementation of IRS and reduced RDT-confirmed all-ages routinely reported malaria case incidence. As expected, the observed effect decreased (while remaining significant) from the first six months post-IRS to the seventh through twelfth months post-IRS, as the residual efficacy of IRS insecticides waned.[40]

In one district, Iakora, IRS did not appear to significantly reduce case incidence compared to if it had not been deployed. Iakora, which received a single IRS campaign in 2020, had the least amount of data (6 out of 11 health facilities and 55% of facility-months) eligible for inclusion in the analysis, which may have affected the model’s ability to detect a significant IRS impact.

When investigating the impact of multiple years of exposure to IRS, we found that three years of exposure produced significantly greater impact than the first or second year. Sustained implementation of IRS in Eastern Uganda was also associated with a significant and sustained reduction in malaria after 4 to 5 years,[3] though there is recent preliminary evidence of a resurgence in the sixth and seventh years of implementation in the same area, which coincided with a shift in insecticide formulation from pirimiphos-methyl to clothianidin.[43] These results suggest that policymakers should consider sustaining implementation of IRS for at least 3 years in target geographies, and give further weight to the tradeoff between the increased benefits of continued IRS versus the potential for a resurgence in malaria cases if IRS is withdrawn, as has been observed in Mali and Uganda.[19,43]

The results of investigating the impact of level of IRS spray coverage were not robust to sensitivity analysis. The extreme skewness in the data (87.9% of communes achieved coverage greater than 90% across all spray campaigns) and lack of variation in coverage levels likely hindered this analysis. Additional studies should be undertaken in contexts where there is greater variation in IRS spray coverage. The results of such a study would be particularly relevant to NMCPs seeking to save costs if levels of coverage below 85% (WHO guideline target) are found to achieve similar impact as high coverage. High IRS coverage (>80%) in Bioko Island, Equatorial Guinea, was demonstrated to offer community protection to individuals in sprayed and unsprayed houses, compared with individuals in communities with <20% coverage,[2] although differences in effect at other levels have not been previously investigated.

The IRS interventions in this study were implemented in communities where mass ITN distributions also took place. Although there is evidence of the individual efficacy of both interventions, few studies have evaluated their joint effect, and the evidence of their combined effectiveness remains inconclusive.[1,44] WHO has recognized the need to close this evidence gap.[45] This study suggests a significant benefit of non-pyrethroid IRS implementation in the presence of standard pyrethroid ITNs on malaria case incidence in Madagascar.

Important limitations of this study include its observational design and reliance on routine passive surveillance data, which was used to generate the primary outcome variable. These data do not account for care-seeking challenges such as patient access to health care, which are likely disproportionately present in rural areas, nor potential differential data reporting consistency among health facilities. It has been estimated that as many as four out of every five malaria cases go uncounted in Madagascar,[46] which would mean that the true malaria cases were undercounted and likely an underestimated number of cases averted at the community level.

However, the availability of routine surveillance data for all years and districts allows us to estimate district-level impact at a much lower cost than collecting data through a survey. Conducted routinely, this type of study is ideal for informing national policy decisions, such as the targeting of vector control interventions in line with the WHO strategy for subnational targeting.[45]

The selection of appropriate controls conducted via propensity score matching represents another limitation related to the observational nature of this study. Although covariate distributional balance between matched IRS and control facilities was ensured, this method cannot truly replicate the benefits of a controlled randomized experiment, nor can it account for the influence of unobserved covariates on the outcome measure.

With regard to the impact of IRS spray coverage on malaria incidence, potential biases include bias of IRS spray teams to report high coverage, although internal post-spray data quality audits performed within 90 days of spray completion help ensure accurate coverage reporting.[25–29] Additionally, there is potential correlation between low spray coverage and difficult-to-access areas where there may also be less access to health care and less likelihood of obtaining RDT confirmation of cases. These potential biases would be expected to influence our findings toward the null effect of no impact of level of IRS coverage on malaria incidence.

## CONCLUSION

The results presented here, interpreted with the limitations described above, suggest a positive public health impact of IRS that bolsters the evidence base around effectiveness of non-pyrethroid insecticides. These results suggest that sustained non-pyrethroid IRS implementation for 3 years may confer additional vector control benefits. This work highlights the value of routine surveillance data for evaluating intervention impact, which empowers evidence-based decision-making as National Malaria Programs seek to implement targeted vector control at subnational levels.

## Data Availability

All data are available from the authors upon reasonable request.

## DISCLAIMER

The findings and conclusions in this report are those of the authors and do not necessarily represent the official position of the U.S. Centers for Disease Control and Prevention and U.S. Agency for International Development.

## CONTRIBUTORSHIP STATEMENT

EH performed descriptive and statistical analysis with the support of JR and SB. SR and HR oversaw programmatic IRS activities including deployment. EH drafted the manuscript with input from HR. AB, LK, SZ, CD, TG, DJ, and CR critically reviewed manuscript drafts and provided feedback. All authors read and approved the final manuscript.

## AVAILABILITY OF DATA AND MATERIALS

All data are available from the authors upon reasonable request.

