## Supplement for "Evaluating the impact of indoor residual spraying on malaria transmission in Madagascar using routine health data"

**SUPPLEMENTAL INFORMATION**

### 1. Health facilities included in the study

Health facility data for all districts was obtained from the Madagascar DHIS2 for the period July 2016 to June 2021 and assessed for inclusion eligibility based on the following criteria:

1. Facilities in districts where IRS was implemented before 2017.
2. Non-public facilities (Hospitals and facilities owned/run by private/religious/NGO entities were excluded due to catchment population overlap with public primary facilities).
3. Facilities missing catchment population data in DHIS2.
4. Facilities in communes missing geocoordinates, which did not allow them to be linked to climate data.
5. Facilities missing RDT-confirmed case data for an entire year.

Following selection of facilities based on the above criteria, the remaining facilities were included in propensity score analysis (described in the next section) where facilities in control districts were matched to facilities in IRS districts. Unmatched control facilities were dropped from the analysis, and the remaining facilities were included in the modeling datasets used in study questions 1 and 2. The modeling dataset for study question 3 retained all IRS facilities and dropped all control facilities (Supplemental Figure 1).

*Supplemental Figure 1. Health facility eligibility and inclusion in the study.*

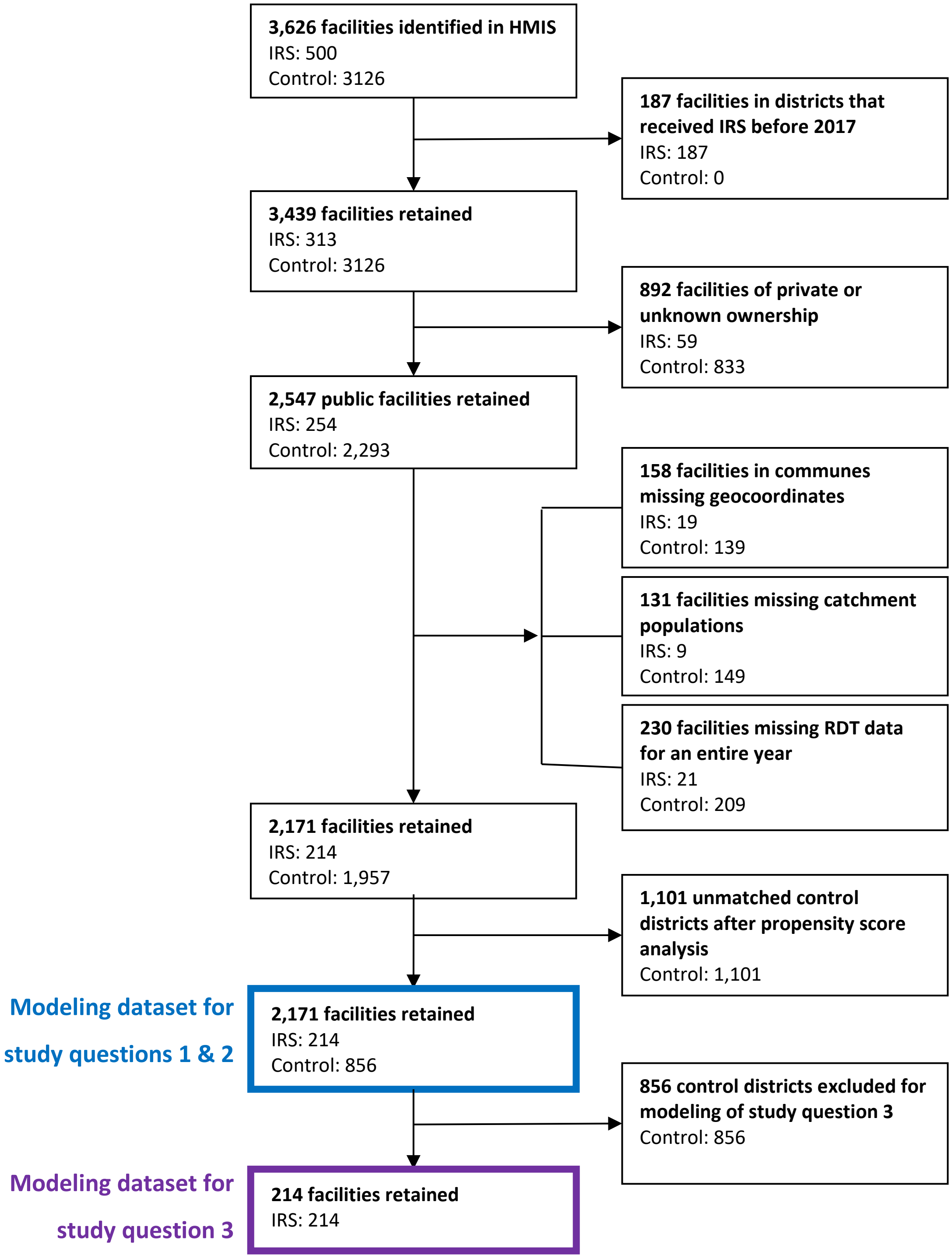

### 2. Propensity score analysis

Propensity score analysis is a method of matching untreated subjects to treated subjects based on even distribution of selected covariates.[1] Broadly, the steps for conducting propensity score analysis are:

1. Select the set of covariates to include.
2. Use logistic regression to obtain a propensity score for each subject.
3. Match exposed and unexposed subjects based on the propensity score.
4. Inspect the balance of covariates in exposed and unexposed groups after matching.

For this analysis, matching was performed at the health facility level using data from the first 12 months of the study period, (July 2016 to June 2017), before IRS had been implemented in any of the study districts. The selected covariates were: EVI, precipitation, ITN survival, and RDT-confirmed all ages malaria case incidence per 1,000 population. Covariates were averaged for each health facility over the 12-month period. Exposure to IRS was the binary outcome variable, where facilities where IRS would be implemented were assigned a 1, and control facilities were assigned a 0.

Control facilities were matched to IRS facilities based on nearest neighbor matching of propensity scores, with a 1:4 ratio of IRS to control. Each control facility could be matched to only one IRS facility.

A total of 856 control facilities were matched to 214 IRS facilities. Match quality was assessed based on comparisons of covariate distributions in control and IRS groups. For continuous covariates, the standard mean difference (SMD) was compared between groups for a threshold of 0.25. The SMD is calculated as:

$$SMD\ of\ X = \frac{\overline{X_1} - \overline{X_2}}{\sqrt{(Var_1 + Var_2)/2}}$$

Where $\overline{X_1}$ and $\overline{X_2}$ are the sample means for the exposed and unexposed groups, respectively, and $Var_1$ and $Var_2$ are sample variances for the exposed and unexposed groups.[2]

Diagnostic plots of matched IRS and control facilities are presented below.

***Supplemental Table 1. Number of IRS and control facilities matched via propensity score matching, per IRS study district.***

| IRS district | IRS | Control |
|---|---|---|
| Ampanihy Ouest | 23 | 92 |
| Betioky Atsimo | 26 | 104 |
| Iakora | 6 | 24 |
| Ihosy | 16 | 64 |
| Manakara Atsimo | 43 | 172 |
| Mananjary | 36 | 144 |
| Sakaraha | 12 | 48 |
| Toliara II | 33 | 132 |
| Vondrozo | 19 | 76 |
| **Total** | **214** | **856** |

***Supplemental Figure 2. Distribution of propensity scores for matched and unmatched facilities using 1:4 nearest neighbor matching without replacement on propensity score. Each circle represents one facility.***

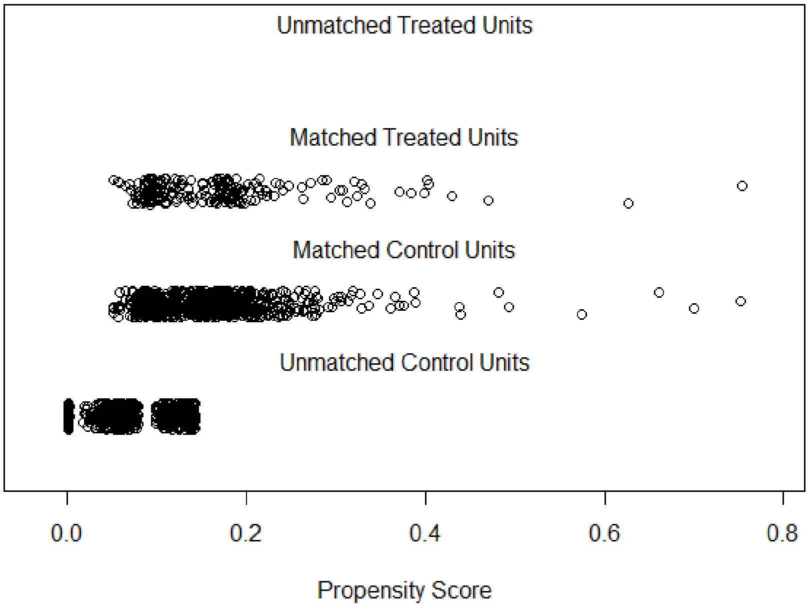

***Supplemental Figure 3. Standardized mean differences (SMD) of covariates before and after matching. Vertical dashed lines represent the 0.25 acceptable threshold.***

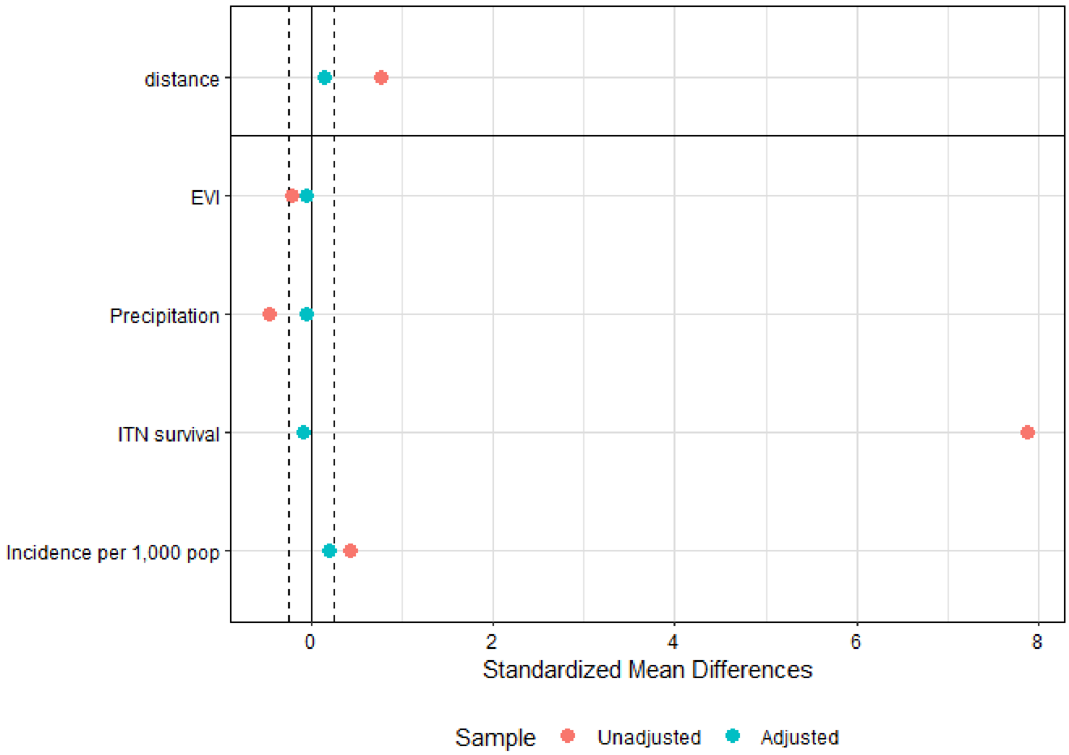

***Supplemental Figure 4. Distributional balances of covariates used in propensity score analysis before and after matching.***

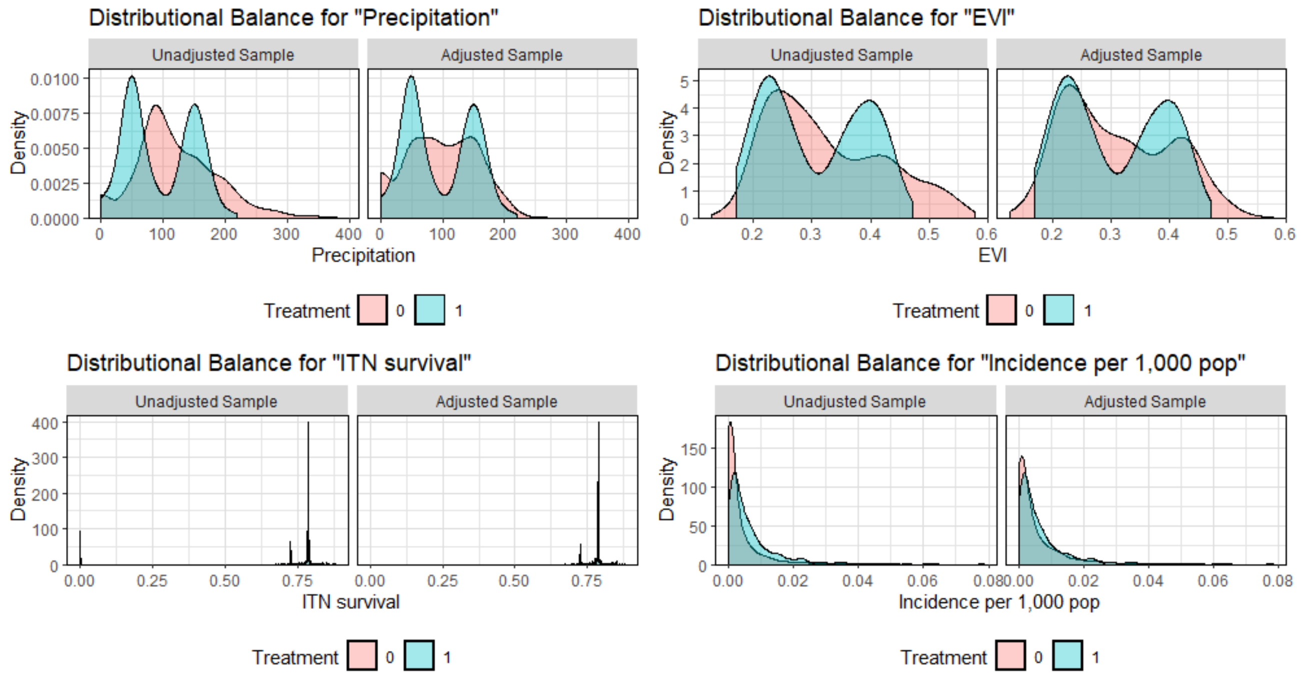

### 3. Madagascar ecozones

To account for varying climate and transmission patterns across Madagascar, precipitation and enhanced vegetation index (EVI) were tested to determine the most appropriate lags for each district. Monthly precipitation and EVI were averaged over the eight malaria transmission ecozones identified by Howes et al.[3] and Pearson correlation tests were performed against monthly reported confirmed malaria cases. The selected lags, as well as a map of the eight ecozones are presented below.

***Supplemental Figure 5. Precipitation and enhanced vegetation index (EVI) lags most correlated with malaria cases in the eight distinct ecozones.***

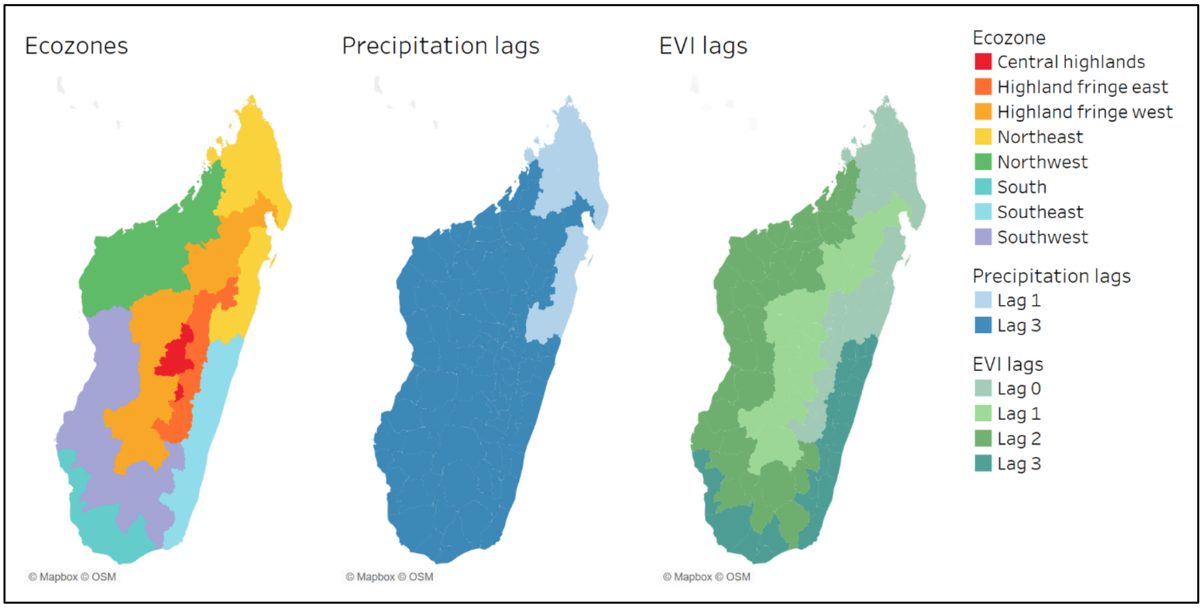

### 4. Coding of IRS exposure covariates

***Supplemental Table 2. Sample table demonstrating how IRS exposure covariates in this study were coded. In this example, IRS was deployed in District A for the first time in October 2019. Blue columns were the exposure covariates included in the model for study question 1; red columns for study question 2; and green columns for study question 3.***

| District | Commune | Health facility | Year | Month | IRS status 0-6 months | IRS status 7-12 months | 12 months post-IRS Year 1 | 12 months post-IRS Year 2 | 12 months post-IRS Year 3 | IRS spray coverage <85% | IRS spray coverage 85-90% | IRS spray coverage 91-95% | IRS spray coverage 96-100% | IRS spray coverage – continuous | RDT-confirmed malaria cases | Other covariates… |
|---|---|---|---|---|---|---|---|---|---|---|---|---|---|---|---|---|
| A | A.1 | A.1.1 | 2019 | Jul | 0 | 0 | 0 | 0 | 0 | 0 | 0 | 0 | 0 | 0 | 16 | |
| A | A.1 | A.1.1 | 2019 | Aug | 0 | 0 | 0 | 0 | 0 | 0 | 0 | 0 | 0 | 0 | 10 | |
| A | A.1 | A.1.1 | 2019 | Sep | 0 | 0 | 0 | 0 | 0 | 0 | 0 | 0 | 0 | 0 | 11 | |
| A | A.1 | A.1.1 | 2019 | Oct | 1 | 0 | 1 | 0 | 0 | 1 | 0 | 0 | 0 | 75% | 42 | |
| A | A.1 | A.1.1 | 2019 | Nov | 1 | 0 | 1 | 0 | 0 | 1 | 0 | 0 | 0 | 75% | 110 | |
| A | A.1 | A.1.1 | 2019 | Dec | 1 | 0 | 1 | 0 | 0 | 1 | 0 | 0 | 0 | 75% | 31 | |
| A | A.1 | A.1.1 | 2020 | Jan | 1 | 0 | 1 | 0 | 0 | 1 | 0 | 0 | 0 | 75% | 136 | |
| A | A.1 | A.1.1 | 2020 | Feb | 1 | 0 | 1 | 0 | 0 | 1 | 0 | 0 | 0 | 75% | 39 | |
| A | A.1 | A.1.1 | 2020 | Mar | 1 | 0 | 1 | 0 | 0 | 1 | 0 | 0 | 0 | 75% | 62 | |
| A | A.1 | A.1.1 | 2020 | Apr | 0 | 1 | 1 | 0 | 0 | 1 | 0 | 0 | 0 | 75% | 29 | |
| A | A.1 | A.1.1 | 2020 | May | 0 | 1 | 1 | 0 | 0 | 1 | 0 | 0 | 0 | 75% | 53 | |
| A | A.1 | A.1.1 | 2020 | Jun | 0 | 1 | 1 | 0 | 0 | 1 | 0 | 0 | 0 | 75% | 67 | |
| A | A.1 | A.1.1 | 2020 | Jul | 0 | 1 | 1 | 0 | 0 | 1 | 0 | 0 | 0 | 75% | 56 | |
| A | A.1 | A.1.1 | 2020 | Aug | 0 | 1 | 1 | 0 | 0 | 1 | 0 | 0 | 0 | 75% | 17 | |
| A | A.1 | A.1.1 | 2020 | Sep | 0 | 1 | 1 | 0 | 0 | 1 | 0 | 0 | 0 | 75% | 38 | |
| A | A.1 | A.1.1 | 2020 | Oct | 0 | 0 | 0 | 0 | 0 | 0 | 0 | 0 | 0 | 0 | 19 | |
| A | A.1 | A.1.1 | 2020 | Nov | 0 | 0 | 0 | 0 | 0 | 0 | 0 | 0 | 0 | 0 | 14 | |
| A | A.1 | A.1.1 | 2020 | Dec | 0 | 0 | 0 | 0 | 0 | 0 | 0 | 0 | 0 | 0 | 51 | |
| A | A.2 | A.2.1 | 2019 | Jul | 0 | 0 | 0 | 0 | 0 | 0 | 0 | 0 | 0 | 0 | 29 | |
| A | A.2 | A.2.1 | 2019 | Aug | 0 | 0 | 0 | 0 | 0 | 0 | 0 | 0 | 0 | 0 | 53 | |
| A | A.2 | A.2.1 | 2019 | Sep | 0 | 0 | 0 | 0 | 0 | 0 | 0 | 0 | 0 | 0 | 57 | |
| A | A.2 | A.2.1 | 2019 | Oct | 1 | 0 | 1 | 0 | 0 | 0 | 0 | 1 | 0 | 93% | 66 | |
| A | A.2 | A.2.1 | 2019 | Nov | 1 | 0 | 1 | 0 | 0 | 0 | 0 | 1 | 0 | 93% | 54 | |
| A | A.2 | A.2.1 | 2019 | Dec | 1 | 0 | 1 | 0 | 0 | 0 | 0 | 1 | 0 | 93% | 32 | |
| A | A.2 | A.2.1 | 2020 | Jan | 1 | 0 | 1 | 0 | 0 | 0 | 0 | 1 | 0 | 93% | 33 | |
| A | A.2 | A.2.1 | 2020 | Feb | 1 | 0 | 1 | 0 | 0 | 0 | 0 | 1 | 0 | 93% | 11 | |
| A | A.2 | A.2.1 | 2020 | Mar | 1 | 0 | 1 | 0 | 0 | 0 | 0 | 1 | 0 | 93% | 19 | |
| A | A.2 | A.2.1 | 2020 | Apr | 0 | 1 | 1 | 0 | 0 | 0 | 0 | 1 | 0 | 93% | 39 | |
| A | A.2 | A.2.1 | 2020 | May | 0 | 1 | 1 | 0 | 0 | 0 | 0 | 1 | 0 | 93% | 33 | |
| A | A.2 | A.2.1 | 2020 | Jun | 0 | 1 | 1 | 0 | 0 | 0 | 0 | 1 | 0 | 93% | 35 | |

| | | | | | | | | | | | | | | | |
|---|---|---|---|---|---|---|---|---|---|---|---|---|---|---|---|
| A | A.2 | A.2.1 | 2020 | Jul | 0 | 1 | 1 | 0 | 0 | 0 | 0 | 1 | 0 | 93% | 8 |
| A | A.2 | A.2.1 | 2020 | Aug | 0 | 1 | 1 | 0 | 0 | 0 | 0 | 1 | 0 | 93% | 4 |
| A | A.2 | A.2.1 | 2020 | Sep | 0 | 1 | 1 | 0 | 0 | 0 | 0 | 1 | 0 | 93% | 4 |
| A | A.2 | A.2.1 | 2020 | Oct | 0 | 0 | 0 | 0 | 0 | 0 | 0 | 0 | 0 | 0 | 0 |
| A | A.2 | A.2.1 | 2020 | Nov | 0 | 0 | 0 | 0 | 0 | 0 | 0 | 0 | 0 | 0 | 1 |
| A | A.2 | A.2.1 | 2020 | Dec | 0 | 0 | 0 | 0 | 0 | 0 | 0 | 0 | 0 | 0 | 2 |

### 5. Summary characteristics of study facilities

***Supplemental Table 3. Summary characteristics and indoor residual spraying (IRS) campaign results in the 9 IRS districts and their matched controls (matching performed by propensity score analysis) for the period July 2016 to June 2021.***

| District | | Number of health facilities | Transmission year* | All ages RDT-confirmed malaria cases | Population | Avg. EVI | Precipitation (mm) | Avg. MDA coverage | Avg. ITN survival | Dates of IRS campaign | IRS product deployed | Population protected by IRS | Structures sprayed | Structures found | District-level spray coverage |
|---|---|---|---|---|---|---|---|---|---|---|---|---|---|---|---|
| Ampanihy Ouest | IRS | 23 | 2016-2017 | 2,733 | 299,785 | 0.2 | 12,217 | | 0.78 | | | | | | |
| | | | 2017-2018 | 4,219 | 317,587 | 0.19 | 10,679 | | 0.49 | | | | | | |
| | | | 2018-2019 | 4,856 | 333,469 | 0.19 | 16,809 | | 0.87 | | | | | | |
| | | | 2019-2020 | 8,538 | 375,958 | 0.22 | 10,102 | | 0.72 | Oct – Dec 2019 | Pirimiphos-methyl | 570,846 | 130,446 | 135,676 | 97.0% |
| | | | 2020-2021 | 10,542 | 408,889 | 0.18 | 11,123 | 90.6% | 0.43 | | | | | | |
| | Control | 92 | 2016-2017 | 23,232 | 763,466 | 0.28 | 74,113 | | 0.79 | | | | | | |
| | | | 2017-2018 | 30,731 | 816,829 | 0.29 | 108,467 | | 0.48 | | | | | | |
| | | | 2018-2019 | 37,652 | 906,646 | 0.28 | 113,490 | | 0.87 | | | | | | |
| | | | 2019-2020 | 76,701 | 931,780 | 0.29 | 97,792 | | 0.72 | | | | | | |
| | | | 2020-2021 | 110,551 | 959,594 | 0.27 | 95,816 | 31.8% | 0.43 | | | | | | |
| Betioky Atsimo | IRS | 26 | 2016-2017 | 15,420 | 196,792 | 0.23 | 14,473 | | 0.78 | | | | | | |
| | | | 2017-2018 | 16,564 | 214,160 | 0.21 | 12,370 | | 0.48 | | | | | | |
| | | | 2018-2019 | 18,265 | 229,151 | 0.22 | 19,799 | | 0.87 | | | | | | |
| | | | 2019-2020 | 15,904 | 257,062 | 0.24 | 12,748 | | 0.72 | Oct – Dec 2019 | Pirimiphos-methyl | 273,856 | 66,590 | 69,679 | 96.4% |
| | | | 2020-2021 | 13,789 | 284,309 | 0.21 | 13,957 | | 0.43 | Nov – Dec 2020 | Pirimiphos-methyl | 285,406 | 69,511 | 71,383 | 97.9% |
| | Control | 104 | 2016-2017 | 40,946 | 754,142 | 0.27 | 60,719 | | 0.79 | | | | | | |
| | | | 2017-2018 | 63,871 | 809,501 | 0.28 | 83,793 | | 0.48 | | | | | | |
| | | | 2018-2019 | 54,894 | 925,836 | 0.27 | 93,620 | | 0.87 | | | | | | |
| | | | 2019-2020 | 98,799 | 954,368 | 0.28 | 80,156 | | 0.72 | | | | | | |
| | | | 2020-2021 | 158,792 | 975,177 | 0.26 | 103,276 | 30.9% | 0.43 | | | | | | |
| Iakora | IRS | 6 | 2016-2017 | 2,261 | 30,151 | 0.28 | 4,420 | | 0.79 | | | | | | |
| | | | 2017-2018 | 3,118 | 30,436 | 0.29 | 6,477 | | 0.46 | | | | | | |

| | | | | | | | | | | | | | | | |
|---|---|---|---|---|---|---|---|---|---|---|---|---|---|---|---|
| | | | 2018-2019 | 9,325 | 39,390 | 0.29 | 8,950 | | 0.87 | | | | | | |
| | | | 2019-2020 | 11,547 | 42,567 | 0.28 | 5,976 | | 0.72 | | | | | | |
| | | | 2020-2021 | 16,896 | 45,447 | 0.27 | 6,109 | | 0.43 | Nov 2020 | Clothianidin/ deltamethrin | 53,137 | 11,698 | 11,981 | 97.6% |
| | Control | 24 | 2016-2017 | 10,054 | 229,034 | 0.27 | 22,640 | | 0.78 | | | | | | |
| | | | 2017-2018 | 15,781 | 251,335 | 0.27 | 30,443 | | 0.49 | | | | | | |
| | | | 2018-2019 | 17,180 | 268,404 | 0.26 | 30,321 | | 0.87 | | | | | | |
| | | | 2019-2020 | 26,108 | 276,315 | 0.27 | 27,547 | | 0.72 | | | | | | |
| | | | 2020-2021 | 35,865 | 284,982 | 0.25 | 27,164 | 20.4% | 0.43 | | | | | | |
| Ihosy | IRS | 16 | 2016-2017 | 13,213 | 147,287 | 0.22 | 10,535 | | 0.78 | | | | | | |
| | | | 2017-2018 | 30,164 | 173,002 | 0.2 | 10,991 | | 0.49 | | | | | | |
| | | | 2018-2019 | 16,822 | 201,560 | 0.21 | 14,853 | | 0.87 | | | | | | |
| | | | 2019-2020 | 18,173 | 237,910 | 0.19 | 10,571 | | 0.72 | Oct – Nov 2019 | Pirimiphos-methyl & Clothianidin/ deltamethrin | 200,563 | 42,911 | 44,617 | 96.5% |
| | | | 2020-2021 | 27,131 | 245,875 | 0.18 | 11,286 | | 0.43 | Nov – Dec 2020 | Clothianidin/ deltamethrin | 202,017 | 43,915 | 44,948 | 98.1% |
| | Control | 64 | 2016-2017 | 17,708 | 502,458 | 0.26 | 47,236 | | 0.79 | | | | | | |
| | | | 2017-2018 | 28,469 | 530,715 | 0.26 | 72,231 | | 0.48 | | | | | | |
| | | | 2018-2019 | 31,601 | 576,840 | 0.27 | 74,462 | | 0.87 | | | | | | |
| | | | 2019-2020 | 40,093 | 589,089 | 0.27 | 64,590 | | 0.72 | | | | | | |
| | | | 2020-2021 | 55,399 | 602,728 | 0.26 | 65,882 | 25.5% | 0.43 | | | | | | |
| Manakara Atsimo | IRS | 43 | 2016-2017 | 10,256 | 324,215 | 0.39 | 74,529 | | 0.79 | | | | | | |
| | | | 2017-2018 | 14,242 | 325,447 | 0.42 | 117,933 | | 0.49 | Jul 2017 | Pirimiphos-methyl | 315,258 | 72,450 | 78,928 | 92.4% |
| | | | 2018-2019 | 14,599 | 357,795 | 0.44 | 125,845 | | 0.87 | Jul 2018 | Pirimiphos-methyl | 345,104 | 76,379 | 86,314 | 89.1% |
| | | | 2019-2020 | 26,058 | 356,185 | 0.44 | 109,918 | | 0.72 | | | | | | |
| | | | 2020-2021 | 27,178 | 357,977 | 0.42 | 82,941 | | 0.43 | | | | | | |
| | Control | 172 | 2016-2017 | 39,099 | 1,261,860 | 0.37 | 247,503 | | 0.78 | | | | | | |
| | | | 2017-2018 | 56,640 | 1,408,655 | 0.37 | 362,453 | | 0.49 | | | | | | |
| | | | 2018-2019 | 58,155 | 1,486,693 | 0.37 | 343,680 | | 0.87 | | | | | | |
| | | | 2019-2020 | 111,292 | 1,452,343 | 0.38 | 314,687 | | 0.72 | | | | | | |
| | | | 2020-2021 | 174,141 | 1,432,328 | 0.37 | 276,288 | 8.5% | 0.43 | | | | | | |
| Mananjary | IRS | 36 | 2016-2017 | 10,515 | 358,748 | 0.39 | 61,225 | | 0.78 | | | | | | |
| | | | 2017-2018 | 8,546 | 387,035 | 0.43 | 97,477 | | 0.49 | Jul 2017 | Pirimiphos-methyl | 249,597 | 58,464 | 62,074 | 94.3% |
| | | | 2018-2019 | 13,070 | 388,691 | 0.43 | 96,145 | | 0.87 | Jul 2018 | Pirimiphos-methyl | 279,822 | 65,245 | 69,561 | 94.2% |

| | | | | | | | | | | | | | | | |
|---|---|---|---|---|---|---|---|---|---|---|---|---|---|---|---|
| | | | 2019-2020 | 22,851 | 322,869 | 0.44 | 95,676 | | 0.72 | | | | | | |
| | | | 2020-2021 | 31,354 | 268,254 | 0.42 | 68,614 | | 0.43 | | | | | | |
| | | | 2016-2017 | 29,699 | 996,159 | 0.38 | 218,581 | | 0.78 | | | | | | |
| | | | 2017-2018 | 46,361 | 1,132,622 | 0.38 | 319,881 | | 0.49 | | | | | | |
| | Control | 144 | 2018-2019 | 49,577 | 1,178,813 | 0.38 | 296,602 | | 0.87 | | | | | | |
| | | | 2019-2020 | 84,464 | 1,153,090 | 0.39 | 278,182 | | 0.71 | | | | | | |
| | | | 2020-2021 | 140,715 | 1,126,693 | 0.37 | 241,959 | 4.4% | 0.43 | | | | | | |
| | | | 2016-2017 | 16,324 | 110,769 | 0.22 | 6,710 | | 0.79 | | | | | | |
| | | | 2017-2018 | 13,302 | 117,650 | 0.2 | 6,181 | | 0.49 | | | | | | |
| | IRS | 12 | 2018-2019 | 12,690 | 127,361 | 0.21 | 9,757 | | 0.87 | Sep 2018 | Pirimiphos-methyl | 128,419 | 30,857 | 32,450 | 94.3% |
| | | | 2019-2020 | 18,844 | 142,683 | 0.22 | 7,123 | | 0.72 | Oct – Nov 2019 | Clothianidin | 110,455 | 27,070 | 27,486 | 98.3% |
| Sakaraha | | | 2020-2021 | 22,694 | 156,968 | 0.21 | 8,136 | | 0.43 | Nov – Dec 2020 | Clothianidin | 110,557 | 26,995 | 27,659 | 98.2% |
| | | | 2016-2017 | 24,944 | 238,109 | 0.27 | 23,730 | | 0.79 | | | | | | |
| | | | 2017-2018 | 26,400 | 253,505 | 0.29 | 40,745 | | 0.48 | | | | | | |
| | Control | 48 | 2018-2019 | 30,471 | 308,787 | 0.29 | 44,361 | | 0.87 | | | | | | |
| | | | 2019-2020 | 50,125 | 316,084 | 0.29 | 37,436 | | 0.72 | | | | | | |
| | | | 2020-2021 | 73,375 | 320,232 | 0.27 | 47,264 | 32.7% | 0.43 | | | | | | |
| | | | 2016-2017 | 22,032 | 193,123 | 0.26 | 12,176 | | 0.77 | | | | | | |
| | | | 2017-2018 | 17,596 | 234,259 | 0.21 | 9,166 | | 0.49 | | | | | | |
| | IRS | 33 | 2018-2019 | 10,804 | 243,733 | 0.24 | 15,687 | | 0.87 | Jul 2018 | Pirimiphos-methyl | 402,343 | 96,335 | 110,118 | 86.5% |
| | | | 2019-2020 | 19,936 | 281,781 | 0.25 | 10,926 | | 0.72 | Oct – Dec 2019 | Clothianidin | 243,637 | 58,937 | 62,930 | 95.2% |
| Toliara II | | | 2020-2021 | 24,741 | 320,085 | 0.22 | 16,156 | | 0.43 | Oct – Dec 2020 | Clothianidin | 256,403 | 63,653 | 65,751 | 97.6% |
| | | | 2016-2017 | 44,413 | 990,562 | 0.27 | 86,060 | | 0.79 | | | | | | |
| | | | 2017-2018 | 59,264 | 1,104,097 | 0.28 | 129,551 | | 0.48 | | | | | | |
| | Control | 132 | 2018-2019 | 64,256 | 1,243,304 | 0.28 | 134,808 | | 0.87 | | | | | | |
| | | | 2019-2020 | 102,508 | 1,302,374 | 0.29 | 116,407 | | 0.72 | | | | | | |
| | | | 2020-2021 | 167,837 | 1,331,847 | 0.27 | 133,577 | 31.7% | 0.43 | | | | | | |
| | | | 2016-2017 | 12,128 | 125,780 | 0.4 | 18,570 | | 0.78 | | | | | | |
| Vondrozo | IRS | 19 | 2017-2018 | 11,424 | 131,842 | 0.41 | 33,188 | | 0.48 | Jul 2017 | Pirimiphos-methyl | 125,374 | 27,690 | 29,203 | 94.8% |
| | | | 2018-2019 | 20,805 | 139,759 | 0.42 | 37,047 | | 0.87 | | | | | | |

| | | | | | | | | |
|---|---|---|---|---|---|---|---|---|
| | | 2019-2020 | 38,724 | 155,179 | 0.43 | 28,605 | | 0.72 |
| | | 2020-2021 | 75,354 | 170,797 | 0.4 | 30,047 | | 0.43 |
| | | 2016-2017 | 19,396 | 577,723 | 0.31 | 76,189 | | 0.78 |
| | | 2017-2018 | 28,432 | 632,720 | 0.31 | 121,053 | | 0.48 |
| Control | 73 | 2018-2019 | 26,927 | 699,788 | 0.31 | 117,264 | | 0.87 |
| | | 2019-2020 | 60,914 | 721,666 | 0.31 | 106,429 | | 0.72 |
| | | 2020-2021 | 93,310 | 739,243 | 0.3 | 101,653 | 14.3% | 0.43 |

#### 6. Statistical results of study question 1: Overall impact of IRS

***Supplemental Table 4. Full regression model coefficients, 95% confidence intervals (95%CI) and P-values describing the association between indoor residual spraying status 0–6 and 7–12 months post-campaign as categorical variables, and RDT-confirmed malaria cases among all ages.***

| | IRR | 95% CI | | P |
|---|---|---|---|---|
| | | Upper | Lower | |
| Cosine(2πt/T) | 1.048 | 1.028 | 1.067 | 0.000 |
| Sine(2πt/T) | 0.653 | 0.583 | 0.732 | 0.000 |
| Transmission year | | | | |
| 2016-2017 | Ref | | | |
| 2017-2018 | 1.096 | 1.016 | 1.183 | 0.018 |
| 2018-2019 | 1.498 | 1.398 | 1.605 | 0.000 |
| 2019-2020 | 1.945 | 1.816 | 2.083 | 0.000 |
| 2020-2021 | 2.286 | 2.112 | 2.474 | 0.000 |
| Transmission year ## Sine(2πt/T) | | | | |
| 2016-2017 | Ref | | | |
| 2017-2018 | 1.586 | 1.511 | 1.664 | 0.000 |
| 2018-2019 | 1.622 | 1.548 | 1.700 | 0.000 |
| 2019-2020 | 1.129 | 1.077 | 1.184 | 0.000 |
| 2020-2021 | 1.423 | 1.358 | 1.492 | 0.000 |
| District | | | | |
| Ambalavao | Ref | | | |
| Ambanja | 0.235 | 0.207 | 0.267 | 0.000 |
| Ambatoboeny | 0.322 | 0.280 | 0.371 | 0.000 |
| Ambatofinandrahana | 0.375 | 0.322 | 0.437 | 0.000 |
| Ambatomainty | 3.550 | 2.949 | 4.274 | 0.000 |
| Ambatondrazaka | 1.427 | 1.265 | 1.611 | 0.000 |
| Ambilobe | 1.007 | 0.837 | 1.211 | 0.945 |
| Amboasary Sud | 0.197 | 0.133 | 0.292 | 0.000 |
| Ambovombe Androy | 0.534 | 0.463 | 0.616 | 0.000 |
| Ampanihy Ouest | 1.692 | 1.288 | 2.222 | 0.000 |
| Amparafaravola | 0.912 | 0.749 | 1.112 | 0.364 |
| Analalava | 0.650 | 0.556 | 0.759 | 0.000 |
| Andapa | 1.070 | 0.940 | 1.217 | 0.306 |
| Andilamena | 1.694 | 1.436 | 1.998 | 0.000 |
| Anjozorobe | 0.190 | 0.163 | 0.223 | 0.000 |
| Ankazoabo Atsimo | 1.898 | 1.685 | 2.139 | 0.000 |
| Ankazobe | 2.151 | 1.870 | 2.473 | 0.000 |

| | | | | |
|---|---|---|---|---|
| Anosibe An'ala | 0.038 | 0.032 | 0.046 | 0.000 |
| Antanambao Manampontsy | 2.532 | 2.098 | 3.057 | 0.000 |
| Antsalova | 0.597 | 0.523 | 0.681 | 0.000 |
| Antsiranana II | 1.013 | 0.885 | 1.160 | 0.847 |
| Antsohihy | 0.132 | 0.114 | 0.154 | 0.000 |
| Bealanana | 0.967 | 0.848 | 1.101 | 0.610 |
| Befandriana Avaratra | 0.299 | 0.203 | 0.439 | 0.000 |
| Befotaka | 1.136 | 0.996 | 1.296 | 0.058 |
| Bekily | 0.338 | 0.262 | 0.436 | 0.000 |
| Belo Sur Tsiribihina | 1.212 | 1.049 | 1.401 | 0.009 |
| Beloha Androy | 1.694 | 1.457 | 1.971 | 0.000 |
| Benenitra | 2.904 | 2.418 | 3.489 | 0.000 |
| Beroroha | 0.423 | 0.368 | 0.488 | 0.000 |
| Besalampy | 0.526 | 0.443 | 0.623 | 0.000 |
| Betafo | 2.432 | 2.060 | 2.871 | 0.000 |
| Betioky Atsimo | 2.247 | 1.952 | 2.586 | 0.000 |
| Betroka | 2.348 | 1.929 | 2.858 | 0.000 |
| Boriziny Port Berge | 1.153 | 0.993 | 1.338 | 0.063 |
| Fenoarivobe | 0.720 | 0.614 | 0.844 | 0.000 |
| Iakora | 0.644 | 0.560 | 0.739 | 0.000 |
| Ifanadiana | 1.150 | 1.025 | 1.291 | 0.017 |
| Ihosy | 0.845 | 0.727 | 0.983 | 0.029 |
| Ikalamavony | 1.700 | 1.444 | 2.002 | 0.000 |
| Ikongo Fort_Carnot | 2.085 | 1.738 | 2.501 | 0.000 |
| Ivohibe | 1.531 | 1.345 | 1.742 | 0.000 |
| Kandreho | 2.552 | 2.048 | 3.181 | 0.000 |
| Maevatanana | 1.355 | 0.909 | 2.018 | 0.136 |
| Mahabo | 0.040 | 0.023 | 0.069 | 0.000 |
| Mahajanga I | 1.145 | 0.985 | 1.330 | 0.078 |
| Mahajanga II | 2.052 | 1.779 | 2.367 | 0.000 |
| Mahanoro | 0.683 | 0.514 | 0.908 | 0.009 |
| Maintirano | 2.920 | 2.555 | 3.336 | 0.000 |
| Mampikony | 0.438 | 0.388 | 0.496 | 0.000 |
| Manakara Atsimo | 0.094 | 0.080 | 0.111 | 0.000 |
| Mananara Avaratra | 3.253 | 2.883 | 3.671 | 0.000 |
| Mananjary | 3.418 | 2.774 | 4.211 | 0.000 |
| Mandoto | 1.192 | 1.055 | 1.347 | 0.005 |
| Mandritsara | 0.757 | 0.646 | 0.887 | 0.001 |
| Manja | 1.059 | 0.935 | 1.199 | 0.366 |
| Maroantsetra | 2.058 | 1.726 | 2.454 | 0.000 |
| Marolambo | 0.057 | 0.049 | 0.067 | 0.000 |
| Marovoay | 4.772 | 3.790 | 6.010 | 0.000 |
| Miandrivazo | 0.185 | 0.162 | 0.213 | 0.000 |

| | | | | |
|---|---|---|---|---|
| Midongy du Sud | 0.271 | 0.237 | 0.310 | 0.000 |
| Mitsinjo | 0.757 | 0.670 | 0.856 | 0.000 |
| Morafenobe | 2.366 | 1.996 | 2.805 | 0.000 |
| Moramanga | 1.646 | 1.443 | 1.879 | 0.000 |
| Morombe | 0.047 | 0.028 | 0.079 | 0.000 |
| Morondava | 0.391 | 0.343 | 0.445 | 0.000 |
| Nosy Be | 1.380 | 1.228 | 1.551 | 0.000 |
| Nosy Boraha Sainte Marie | 2.207 | 1.835 | 2.654 | 0.000 |
| Nosy Varika | 2.387 | 2.031 | 2.806 | 0.000 |
| Sakaraha | 0.751 | 0.604 | 0.935 | 0.010 |
| Sambava | 2.099 | 1.698 | 2.595 | 0.000 |
| Soalala | 0.760 | 0.640 | 0.904 | 0.002 |
| Soanierana Ivongo | 0.798 | 0.707 | 0.902 | 0.000 |
| Taolagnaro | 0.034 | 0.029 | 0.039 | 0.000 |
| Toamasina I | 0.696 | 0.579 | 0.835 | 0.000 |
| Toliara I | 1.832 | 1.541 | 2.177 | 0.000 |
| Toliara II | 0.660 | 0.580 | 0.751 | 0.000 |
| Tsaratanana | 0.216 | 0.187 | 0.251 | 0.000 |
| Tsihombe | 1.771 | 1.556 | 2.015 | 0.000 |
| Tsiroanomandidy | 1.587 | 1.384 | 1.819 | 0.000 |
| Vangaindrano | 0.655 | 0.527 | 0.814 | 0.000 |
| Vatomandry | 1.459 | 1.276 | 1.669 | 0.000 |
| Vavatenina | 1.025 | 0.888 | 1.184 | 0.736 |
| Vohimarina | 1.987 | 1.684 | 2.343 | 0.000 |
| Vondrozo | 0.162 | 0.132 | 0.199 | 0.000 |

District ## Sine(2πt/T)

| | | | | |
|---|---|---|---|---|
| Ambalavao | Ref | | | |
| Ambanja | 0.932 | 0.803 | 1.082 | 0.354 |
| Ambatoboeny | 1.293 | 1.105 | 1.513 | 0.001 |
| Ambatofinandrahana | 0.280 | 0.233 | 0.336 | 0.000 |
| Ambatomainty | 1.045 | 0.846 | 1.291 | 0.683 |
| Ambatondrazaka | 0.634 | 0.548 | 0.733 | 0.000 |
| Ambilobe | 1.039 | 0.835 | 1.292 | 0.734 |
| Amboasary Sud | 1.136 | 0.697 | 1.850 | 0.609 |
| Ambovombe Androy | 0.249 | 0.208 | 0.297 | 0.000 |
| Ampanihy Ouest | 0.514 | 0.358 | 0.737 | 0.000 |
| Amparafaravola | 0.318 | 0.250 | 0.404 | 0.000 |
| Analalava | 0.248 | 0.207 | 0.297 | 0.000 |
| Andapa | 0.572 | 0.497 | 0.659 | 0.000 |
| Andilamena | 0.453 | 0.369 | 0.557 | 0.000 |
| Anjozorobe | 1.225 | 1.028 | 1.460 | 0.023 |
| Ankazoabo Atsimo | 0.532 | 0.464 | 0.611 | 0.000 |

| | | | | |
|---|---|---|---|---|
| Ankazobe | 0.962 | 0.817 | 1.132 | 0.641 |
| Anosibe An'ala | 0.805 | 0.640 | 1.013 | 0.065 |
| Antanambao Manampontsy | 0.912 | 0.729 | 1.141 | 0.421 |
| Antsalova | 0.765 | 0.657 | 0.891 | 0.001 |
| Antsiranana II | 0.391 | 0.337 | 0.453 | 0.000 |
| Antsohihy | 0.236 | 0.193 | 0.289 | 0.000 |
| Bealanana | 2.274 | 1.955 | 2.645 | 0.000 |
| Befandriana Avaratra | 1.304 | 0.806 | 2.110 | 0.280 |
| Befotaka | 1.742 | 1.494 | 2.031 | 0.000 |
| Bekily | 0.951 | 0.711 | 1.271 | 0.733 |
| Belo Sur Tsiribihina | 0.386 | 0.322 | 0.464 | 0.000 |
| Beloha Androy | 0.664 | 0.555 | 0.793 | 0.000 |
| Benenitra | 0.456 | 0.364 | 0.571 | 0.000 |
| Beroroha | 0.324 | 0.276 | 0.381 | 0.000 |
| Besalampy | 1.731 | 1.427 | 2.099 | 0.000 |
| Betafo | 0.908 | 0.744 | 1.107 | 0.339 |
| Betioky Atsimo | 0.771 | 0.655 | 0.908 | 0.002 |
| Betroka | 0.308 | 0.240 | 0.395 | 0.000 |
| Boriziny Port Berge | 0.284 | 0.239 | 0.338 | 0.000 |
| Fenoarivobe | 1.281 | 1.067 | 1.538 | 0.008 |
| Iakora | 0.908 | 0.772 | 1.068 | 0.244 |
| Ifanadiana | 1.304 | 1.139 | 1.494 | 0.000 |
| Ihosy | 1.499 | 1.255 | 1.791 | 0.000 |
| Ikalamavony | 0.420 | 0.344 | 0.512 | 0.000 |
| Ikongo Fort_Carnot | 1.040 | 0.842 | 1.286 | 0.714 |
| Ivohibe | 1.679 | 1.434 | 1.967 | 0.000 |
| Kandreho | 0.398 | 0.296 | 0.533 | 0.000 |
| Maevatanana | 0.910 | 0.549 | 1.507 | 0.713 |
| Mahabo | 0.927 | 0.460 | 1.865 | 0.831 |
| Mahajanga I | 0.904 | 0.761 | 1.075 | 0.253 |
| Mahajanga II | 0.886 | 0.753 | 1.044 | 0.149 |
| Mahanoro | 0.301 | 0.212 | 0.426 | 0.000 |
| Maintirano | 0.616 | 0.529 | 0.718 | 0.000 |
| Mampikony | 1.010 | 0.874 | 1.166 | 0.897 |
| Manakara Atsimo | 1.702 | 1.402 | 2.067 | 0.000 |
| Mananara Avaratra | 0.801 | 0.699 | 0.919 | 0.002 |
| Mananjary | 1.255 | 0.981 | 1.605 | 0.070 |
| Mandoto | 0.646 | 0.566 | 0.737 | 0.000 |
| Mandritsara | 0.369 | 0.311 | 0.437 | 0.000 |
| Manja | 0.477 | 0.417 | 0.545 | 0.000 |
| Maroantsetra | 1.103 | 0.893 | 1.362 | 0.363 |
| Marolambo | 0.760 | 0.623 | 0.926 | 0.007 |
| Marovoay | 0.921 | 0.698 | 1.214 | 0.557 |

| | | | | |
|---|---|---|---|---|
| Miandrivazo | 0.992 | 0.847 | 1.162 | 0.919 |
| Midongy du Sud | 0.198 | 0.167 | 0.234 | 0.000 |
| Mitsinjo | 1.018 | 0.887 | 1.168 | 0.798 |
| Morafenobe | 1.100 | 0.897 | 1.350 | 0.361 |
| Moramanga | 0.428 | 0.366 | 0.500 | 0.000 |
| Morombe | 0.769 | 0.393 | 1.505 | 0.443 |
| Morondava | 0.323 | 0.275 | 0.379 | 0.000 |
| Nosy Be | 1.376 | 1.199 | 1.579 | 0.000 |
| Nosy Boraha Sainte Marie | 0.592 | 0.474 | 0.740 | 0.000 |
| Nosy Varika | 1.581 | 1.305 | 1.915 | 0.000 |
| Sakaraha | 0.643 | 0.506 | 0.817 | 0.000 |
| Sambava | 0.482 | 0.369 | 0.630 | 0.000 |
| Soalala | 1.164 | 0.954 | 1.419 | 0.135 |
| Soanierana Ivongo | 1.088 | 0.943 | 1.256 | 0.249 |
| Taolagnaro | 1.402 | 1.171 | 1.680 | 0.000 |
| Toamasina I | 0.392 | 0.318 | 0.482 | 0.000 |
| Toliara I | 1.048 | 0.858 | 1.281 | 0.645 |
| Toliara II | 0.433 | 0.368 | 0.510 | 0.000 |
| Tsaratanana | 1.434 | 1.208 | 1.702 | 0.000 |
| Tsihombe | 0.801 | 0.687 | 0.933 | 0.004 |
| Tsiroanomandidy | 1.312 | 1.117 | 1.541 | 0.001 |
| Vangaindrano | 0.429 | 0.337 | 0.545 | 0.000 |
| Vatomandry | 0.375 | 0.323 | 0.434 | 0.000 |
| Vavatenina | 0.597 | 0.510 | 0.699 | 0.000 |
| Vohimarina | 1.404 | 1.144 | 1.722 | 0.001 |
| Vondrozo | 0.306 | 0.237 | 0.395 | 0.000 |
| IRS exposure | | | | |
| 0-6 months post-IRS | 0.567 | 0.531 | 0.605 | 0.000 |
| 7-12 months post-IRS | 0.752 | 0.704 | 0.802 | 0.000 |
| Precipitation (lagged and scaled) | 1.025 | 1.010 | 1.041 | 0.001 |
| EVI (lagged and scaled) | 1.143 | 1.113 | 1.174 | 0.000 |
| LLIN survival | 0.365 | 0.326 | 0.408 | 0.000 |
| MDA coverage | 0.796 | 0.699 | 0.908 | 0.001 |
| Observations | 61,828 | | | |
| Number of groups | 214 | | | |

***Supplemental Figure 6. Study question 1 modeled estimates and residuals. (A) Modeled incidence of RDT-confirmed all-ages malaria incidence per 1,000 population in nine IRS districts from July 2016 to June 2017. Vertical dashed lines indicate the dates of IRS campaigns in each district. Solid lines represent modeled estimates and points represent observed values reported in DHIS2. Shading around the solid lines represent 95% confidence intervals of model estimates. (B) Plot of Anscombe's residuals. (B) Plot of Anscombe's residuals by matched cluster.***

**A.**

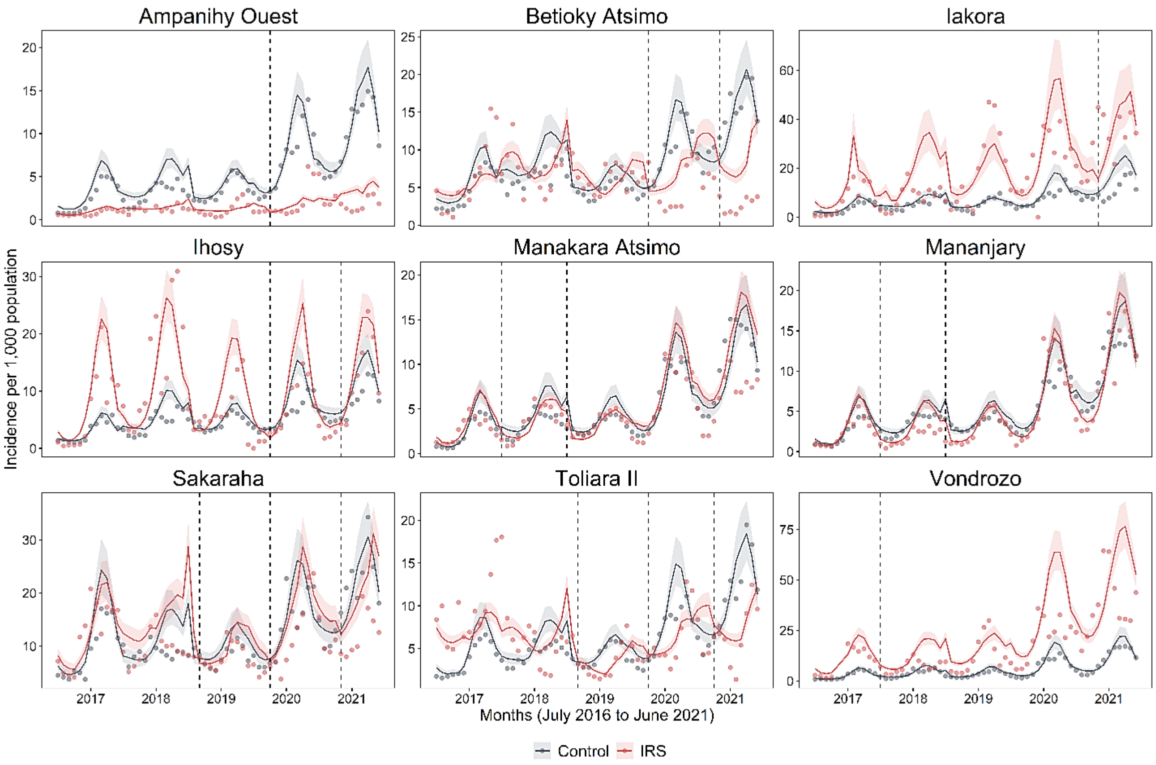

**B.**

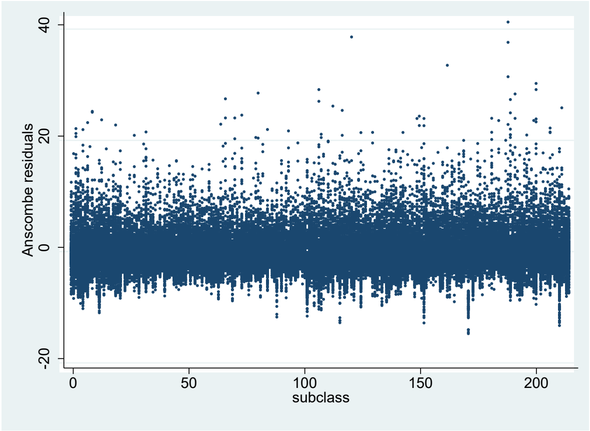

**7. Statistical results of study question 2: Impact of sustained years of exposure to IRS**

***Supplemental Table 5. Full regression model coefficients, 95% confidence intervals (95%CI) and P-values describing the association between indoor residual spraying status in the first, second, or third year of IRS implementation, and RDT-confirmed malaria cases among all ages.***

| | IRR | 95% CI | | P |
|---|---|---|---|---|
| | | Upper | Lower | |
| Transmission year | | | | |
| 2016-2017 | Ref | | | |
| 2017-2018 | 1.194 | 1.140 | 1.251 | 0.000 |
| 2018-2019 | 1.434 | 1.384 | 1.485 | 0.000 |
| 2019-2020 | 1.992 | 1.924 | 2.062 | 0.000 |
| 2020-2021 | 2.636 | 2.503 | 2.776 | 0.000 |
| Month | | | | |
| January | 0.591 | 0.562 | 0.622 | 0.000 |
| February | 0.491 | 0.466 | 0.517 | 0.000 |
| March | 0.460 | 0.437 | 0.485 | 0.000 |
| April | 0.491 | 0.467 | 0.517 | 0.000 |
| May | 0.574 | 0.546 | 0.604 | 0.000 |
| June | 0.706 | 0.672 | 0.743 | 0.000 |
| July | Ref | | | |
| August | 1.048 | 0.997 | 1.100 | 0.064 |
| September | 1.146 | 1.089 | 1.206 | 0.000 |
| October | 1.320 | 1.249 | 1.396 | 0.000 |
| November | 1.361 | 1.289 | 1.436 | 0.000 |
| December | 1.058 | 1.003 | 1.117 | 0.038 |
| District | | | | |
| Ambalavao | Ref | | | |
| Ambanja | 0.383 | 0.334 | 0.439 | 0.000 |
| Ambatoboeny | 0.518 | 0.442 | 0.607 | 0.000 |
| Ambatofinandrahana | 1.750 | 1.463 | 2.093 | 0.000 |
| Ambatomainty | 0.350 | 0.307 | 0.399 | 0.000 |
| Ambatondrazaka | 1.935 | 1.593 | 2.349 | 0.000 |
| Ambilobe | 0.315 | 0.278 | 0.356 | 0.000 |
| Amboasary Sud | 0.272 | 0.240 | 0.309 | 0.000 |
| Ambovombe Androy | 1.580 | 1.380 | 1.808 | 0.000 |
| Ampanihy Ouest | 0.170 | 0.149 | 0.193 | 0.000 |
| Amparafaravola | 0.309 | 0.273 | 0.349 | 0.000 |
| Analalava | 0.533 | 0.468 | 0.606 | 0.000 |
| Andapa | 1.536 | 1.299 | 1.817 | 0.000 |

| | | | | |
|---|---|---|---|---|
| Andilamena | 0.318 | 0.247 | 0.408 | 0.000 |
| Anjozorobe | 0.959 | 0.834 | 1.104 | 0.560 |
| Ankazoabo Atsimo | 0.124 | 0.107 | 0.144 | 0.000 |
| Ankazobe | 1.556 | 1.349 | 1.796 | 0.000 |
| Anosibe An'ala | 0.524 | 0.461 | 0.595 | 0.000 |
| Antanambao Manampontsy | 0.763 | 0.664 | 0.876 | 0.000 |
| Antsalova | 1.030 | 0.858 | 1.236 | 0.751 |
| Antsiranana II | 2.130 | 1.820 | 2.493 | 0.000 |
| Antsohihy | 0.028 | 0.025 | 0.032 | 0.000 |
| Bealanana | 0.870 | 0.753 | 1.005 | 0.058 |
| Befandriana Avaratra | 0.148 | 0.131 | 0.167 | 0.000 |
| Befotaka | 0.616 | 0.556 | 0.682 | 0.000 |
| Bekily | 2.688 | 2.229 | 3.240 | 0.000 |
| Belo Sur Tsiribihina | 0.594 | 0.524 | 0.673 | 0.000 |
| Beloha Androy | 1.188 | 1.044 | 1.353 | 0.009 |
| Benenitra | 0.177 | 0.153 | 0.206 | 0.000 |
| Beroroha | 1.357 | 1.136 | 1.620 | 0.001 |
| Besalampy | 1.247 | 1.096 | 1.419 | 0.001 |
| Betafo | 0.677 | 0.569 | 0.804 | 0.000 |
| Betioky Atsimo | 0.185 | 0.152 | 0.225 | 0.000 |
| Betroka | 1.095 | 0.973 | 1.232 | 0.132 |
| Boriziny Port Berge | 0.981 | 0.864 | 1.114 | 0.766 |
| Fenoarivobe | 0.622 | 0.533 | 0.725 | 0.000 |
| Iakora | 1.456 | 1.103 | 1.922 | 0.008 |
| Ifanadiana | 1.811 | 1.507 | 2.177 | 0.000 |
| Ihosy | 1.300 | 1.141 | 1.481 | 0.000 |
| Ikalamavony | 1.347 | 1.180 | 1.537 | 0.000 |
| Ikongo Fort_Carnot | 1.400 | 1.187 | 1.650 | 0.000 |
| Ivohibe | 1.031 | 0.925 | 1.148 | 0.583 |
| Kandreho | 2.155 | 1.725 | 2.693 | 0.000 |
| Maevatanana | 3.324 | 2.647 | 4.175 | 0.000 |
| Mahabo | 1.809 | 1.537 | 2.131 | 0.000 |
| Mahajanga I | 0.787 | 0.689 | 0.898 | 0.000 |
| Mahajanga II | 0.181 | 0.120 | 0.271 | 0.000 |
| Mahanoro | 1.131 | 0.750 | 1.706 | 0.558 |
| Maintirano | 1.307 | 1.146 | 1.491 | 0.000 |
| Mampikony | 0.745 | 0.618 | 0.900 | 0.002 |
| Manakara Atsimo | 1.137 | 1.034 | 1.250 | 0.008 |
| Mananara Avaratra | 0.709 | 0.577 | 0.871 | 0.001 |
| Mananjary | 1.002 | 0.908 | 1.106 | 0.963 |
| Mandoto | 1.276 | 1.081 | 1.507 | 0.004 |
| Mandritsara | 0.488 | 0.434 | 0.549 | 0.000 |
| Manja | 1.270 | 1.113 | 1.449 | 0.000 |

| | | | | |
|---|---|---|---|---|
| Maroantsetra | 0.720 | 0.583 | 0.890 | 0.002 |
| Marolambo | 1.524 | 1.357 | 1.712 | 0.000 |
| Marovoay | 2.615 | 2.118 | 3.227 | 0.000 |
| Miandrivazo | 0.477 | 0.415 | 0.548 | 0.000 |
| Midongy du Sud | 2.991 | 2.527 | 3.540 | 0.000 |
| Mitsinjo | 1.590 | 1.358 | 1.863 | 0.000 |
| Morafenobe | 1.869 | 1.573 | 2.220 | 0.000 |
| Moramanga | 0.458 | 0.400 | 0.525 | 0.000 |
| Morombe | 0.949 | 0.833 | 1.082 | 0.438 |
| Morondava | 0.800 | 0.691 | 0.926 | 0.003 |
| Nosy Be | 0.273 | 0.184 | 0.405 | 0.000 |
| Nosy Boraha Sainte Marie | 0.044 | 0.027 | 0.074 | 0.000 |
| Nosy Varika | 0.920 | 0.815 | 1.040 | 0.182 |
| Sakaraha | 1.524 | 1.321 | 1.758 | 0.000 |
| Sambava | 0.041 | 0.024 | 0.071 | 0.000 |
| Soalala | 2.700 | 2.247 | 3.245 | 0.000 |
| Soanierana Ivongo | 0.775 | 0.586 | 1.025 | 0.074 |
| Taolagnaro | 1.644 | 1.481 | 1.824 | 0.000 |
| Toamasina I | 0.032 | 0.027 | 0.039 | 0.000 |
| Toliara I | 0.090 | 0.075 | 0.107 | 0.000 |
| Toliara II | 0.973 | 0.869 | 1.090 | 0.636 |
| Tsaratanana | 1.306 | 1.124 | 1.517 | 0.000 |
| Tsihombe | 0.042 | 0.036 | 0.049 | 0.000 |
| Tsiroanomandidy | 1.696 | 1.372 | 2.097 | 0.000 |
| Vangaindrano | 2.764 | 2.498 | 3.058 | 0.000 |
| Vatomandry | 0.791 | 0.690 | 0.907 | 0.001 |
| Vavatenina | 0.752 | 0.633 | 0.892 | 0.001 |
| Vohimarina | 0.166 | 0.144 | 0.192 | 0.000 |
| Vondrozo | 2.509 | 2.235 | 2.818 | 0.000 |
| IRS exposure | | | | |
| Year 1 | 0.629858 | 0.593584 | 0.668348 | 0 |
| Year 2 | 0.597635 | 0.554576 | 0.644038 | 0 |
| Year 3 | 0.422875 | 0.355057 | 0.503647 | 0 |
| Precipitation (lagged and scaled) | 0.980 | 0.964 | 0.995 | 0.012 |
| EVI (lagged and scaled) | 1.001 | 0.977 | 1.026 | 0.939 |
| LLIN survival | 0.532 | 0.477 | 0.593 | 0.000 |
| MDA coverage | 0.632 | 0.552 | 0.724 | 0.000 |
| Observations | 61,828 | | | |
| Number of groups | 214 | | | |

***Supplemental Figure 7. Study question 2 modeled estimates and residuals. (A) Modeled incidence of RDT-confirmed all-ages malaria incidence per 1,000 population in nine IRS districts from July 2016 to June 2017. Vertical dashed lines indicate the dates of IRS campaigns in each district. Solid lines represent modeled estimates and points represent observed values reported in DHIS2. Shading around the solid lines represent 95% confidence intervals of model estimates. (B) Plot of Anscombe's residuals.***

**A.**

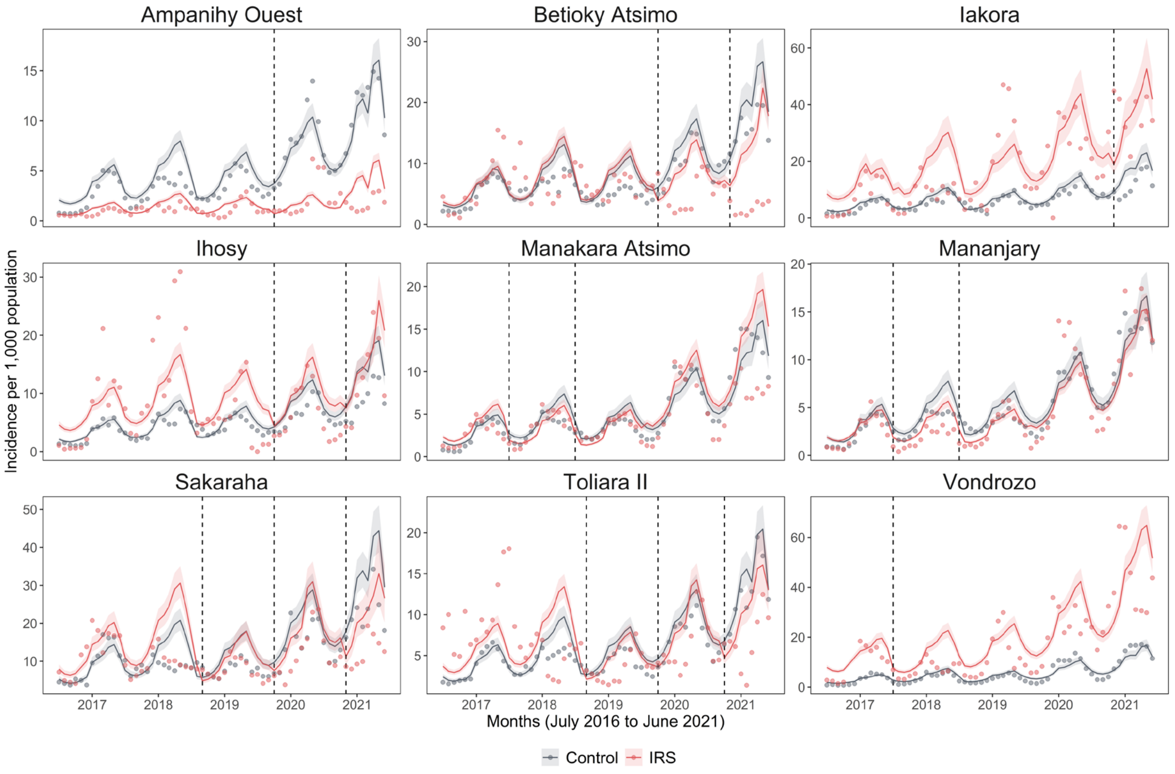

**B.**

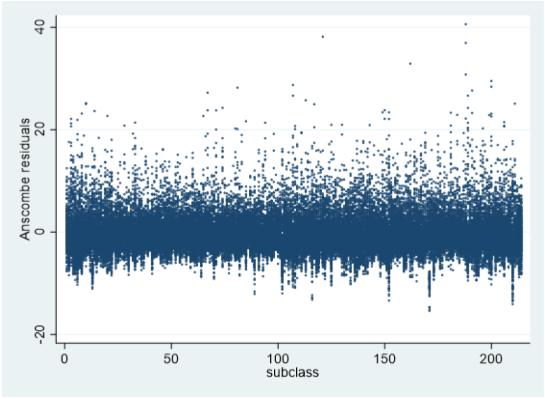

### 8. Statistical results of study question 3: Impact of level of IRS spray coverage

#### 8.1. IRS spray coverage modeled as a categorical variable

***Supplemental Table 6. Full regression model coefficients, 95% confidence intervals (95%CI) and P-values resulting from a model describing the association between RDT-confirmed malaria cases among all ages and IRS spray coverage as a categorical variable with four coverage bins: ≤85%, 86%–90%, 91%–95%, and 96%–100%.***

| | IRR | 95% CI | | P |
|---|---|---|---|---|
| | | Upper | Lower | |
| Cosine(2πt/T) | 1.177 | 1.125 | 1.232 | 0.000 |
| Sine(2πt/T) | 0.429 | 0.378 | 0.487 | 0.000 |
| Transmission year | | | | |
| 2016-2017 | Ref | | | |
| 2017-2018 | 1.294 | 1.174 | 1.426 | 0.000 |
| 2018-2019 | 1.733 | 1.597 | 1.880 | 0.000 |
| 2019-2020 | 2.218 | 1.959 | 2.511 | 0.000 |
| 2020-2021 | 2.162 | 1.858 | 2.517 | 0.000 |
| Transmission year ## Sine(2πt/T) | | | | |
| 2016-2017 | Ref | | | |
| 2017-2018 | 1.859 | 1.661 | 2.080 | 0.000 |
| 2018-2019 | 1.600 | 1.431 | 1.789 | 0.000 |
| 2019-2020 | 1.270 | 1.063 | 1.518 | 0.009 |
| 2020-2021 | 1.314 | 1.091 | 1.582 | 0.004 |
| District | | | | |
| Ampanihy Ouest | Ref | | | |
| Betioky Atsimo | 0.585 | 0.315 | 1.086 | 0.089 |
| Iakora | 2.999 | 1.710 | 5.261 | 0.000 |
| Ihosy | 0.619 | 0.357 | 1.074 | 0.088 |
| Manakara Atsimo | 1.141 | 0.745 | 1.748 | 0.544 |
| Mananjary | 0.640 | 0.383 | 1.069 | 0.088 |
| Sakaraha | 1.044 | 0.594 | 1.836 | 0.881 |
| Toliara II | 2.629 | 1.543 | 4.479 | 0.000 |
| Vondrozo | 12.349 | 3.610 | 42.249 | 0.000 |
| District ## Sine(2πt/T) | | | | |
| Ampanihy Ouest | Ref | | | |
| Betioky Atsimo | 0.533 | 0.451 | 0.630 | 0.000 |
| Iakora | 0.868 | 0.684 | 1.102 | 0.246 |
| Ihosy | 2.054 | 1.604 | 2.631 | 0.000 |

| | | | | |
|---|---|---|---|---|
| Manakara Atsimo | 0.624 | 0.541 | 0.718 | 0.000 |
| Mananjary | 0.577 | 0.491 | 0.678 | 0.000 |
| Sakaraha | 0.629 | 0.544 | 0.727 | 0.000 |
| Toliara II | 0.719 | 0.572 | 0.905 | 0.005 |
| Vondrozo | 2.254 | 1.051 | 4.833 | 0.037 |
| IRS spray coverage | | | | |
| ≤85% | Ref | | | |
| 86-90% | 0.803 | 0.690 | 0.934 | 0.005 |
| 91-95% | 0.940 | 0.830 | 1.064 | 0.329 |
| 96-100% | 1.070 | 0.944 | 1.214 | 0.289 |
| Precipitation (lagged and scaled) | 0.991 | 0.960 | 1.023 | 0.587 |
| EVI (lagged and scaled) | 1.185 | 1.101 | 1.274 | 0.000 |
| LLIN survival | 0.759 | 0.616 | 0.935 | 0.010 |
| Observations | 8,882 | | | |
| Number of groups | 246 | | | |

***Supplemental Figure 7. Study question 3 modeled estimates and residuals – IRS as a categorical variable. (A) Modeled incidence of RDT-confirmed all-ages malaria incidence per 1,000 population in nine IRS districts from July 2016 to June 2017 where IRS spray coverage is modeled as a categorical variable with four coverage bins: ≤85%, 86%–90%, 91%–95%, and 96%–100%. Data was restricted include up to 12 months following each spray campaign Vertical dashed lines indicate the dates of IRS campaigns in each district. Solid lines represent modeled estimates and points represent observed values reported in DHIS2. Shading around the solid lines represent 95% confidence intervals of model estimates. (B) Plot of Anscombe's residuals by commune.***

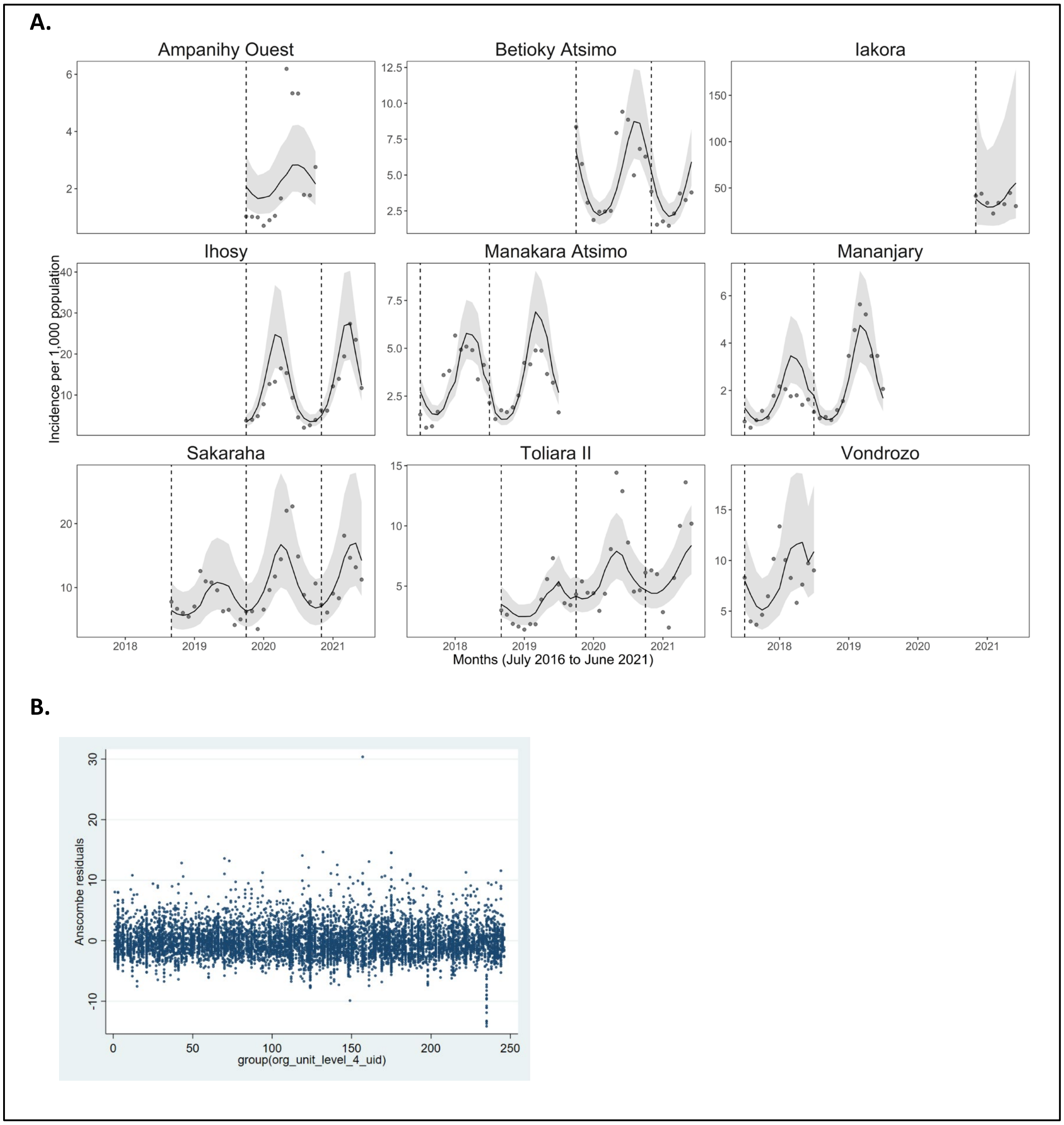

### 8.2. IRS spray coverage modeled as a continuous variable

***Supplemental Table 7. Full regression model coefficients, 95% confidence intervals (95%CI) and P-values resulting from a model describing the association between RDT-confirmed malaria cases among all ages and IRS spray coverage as a continuous variable.***

| | IRR | 95% CI | | P |
|---|---|---|---|---|
| | | Upper | Lower | |
| Cosine(2πt/T) | 1.175 | 1.123 | 1.230 | 0.000 |
| Sine(2πt/T) | 0.430 | 0.378 | 0.488 | 0.000 |
| Transmission year | | | | |
| 2016-2017 | Ref | | | |
| 2017-2018 | 1.302 | 1.182 | 1.435 | 0.000 |
| 2018-2019 | 1.756 | 1.619 | 1.904 | 0.000 |
| 2019-2020 | 2.200 | 1.946 | 2.488 | 0.000 |
| 2020-2021 | 2.186 | 1.881 | 2.540 | 0.000 |
| Transmission year ## Sine(2πt/T) | | | | |
| 2016-2017 | Ref | | | |
| 2017-2018 | 1.864 | 1.665 | 2.086 | 0.000 |
| 2018-2019 | 1.604 | 1.434 | 1.794 | 0.000 |
| 2019-2020 | 1.301 | 1.089 | 1.555 | 0.004 |
| 2020-2021 | 1.313 | 1.090 | 1.582 | 0.004 |
| District | | | | |
| Ampanihy Ouest | Ref | | | |
| Betioky Atsimo | 0.562 | 0.303 | 1.040 | 0.066 |
| Iakora | 2.979 | 1.703 | 5.212 | 0.000 |
| Ihosy | 0.624 | 0.361 | 1.080 | 0.092 |
| Manakara Atsimo | 1.115 | 0.730 | 1.704 | 0.615 |
| Mananjary | 0.622 | 0.373 | 1.036 | 0.068 |
| Sakaraha | 1.017 | 0.580 | 1.782 | 0.954 |
| Toliara II | 2.612 | 1.537 | 4.438 | 0.000 |
| Vondrozo | 12.463 | 3.661 | 42.424 | 0.000 |
| District ## Sine(2πt/T) | | | | |
| Ampanihy Ouest | Ref | | | |
| Betioky Atsimo | 0.531 | 0.449 | 0.628 | 0.000 |
| Iakora | 0.864 | 0.681 | 1.098 | 0.231 |
| Ihosy | 2.003 | 1.564 | 2.565 | 0.000 |
| Manakara Atsimo | 0.620 | 0.538 | 0.714 | 0.000 |
| Mananjary | 0.576 | 0.490 | 0.677 | 0.000 |

| | | | | |
|---|---|---|---|---|
| Sakaraha | 0.626 | 0.541 | 0.723 | 0.000 |
| Toliara II | 0.709 | 0.563 | 0.892 | 0.003 |
| Vondrozo | 2.254 | 1.050 | 4.836 | 0.037 |
| IRS spray coverage | 1.010 | 1.003 | 1.016 | 0.003 |
| Precipitation (lagged and scaled) | 0.992 | 0.961 | 1.024 | 0.615 |
| EVI (lagged and scaled) | 1.189 | 1.105 | 1.279 | 0.000 |
| LLIN survival | 0.747 | 0.607 | 0.921 | 0.006 |
| Observations | 8,882 | | | |
| Number of groups | 246 | | | |

***Supplemental Figure 8. Study question 3 modeled estimates and residuals – IRS as a continuous variable. (A) Modeled incidence of RDT-confirmed all-ages malaria incidence per 1,000 population in nine IRS districts from July 2016 to June 2017 where IRS spray coverage is modeled as a continuous variable. Data was restricted include up to 12 months following each spray campaign Vertical dashed lines indicate the dates of IRS campaigns in each district. Solid lines represent modeled estimates and points represent observed values reported in DHIS2. Shading around the solid lines represent 95% confidence intervals of model estimates. (B) Plot of Anscombe's residuals by commune.***

**A.**

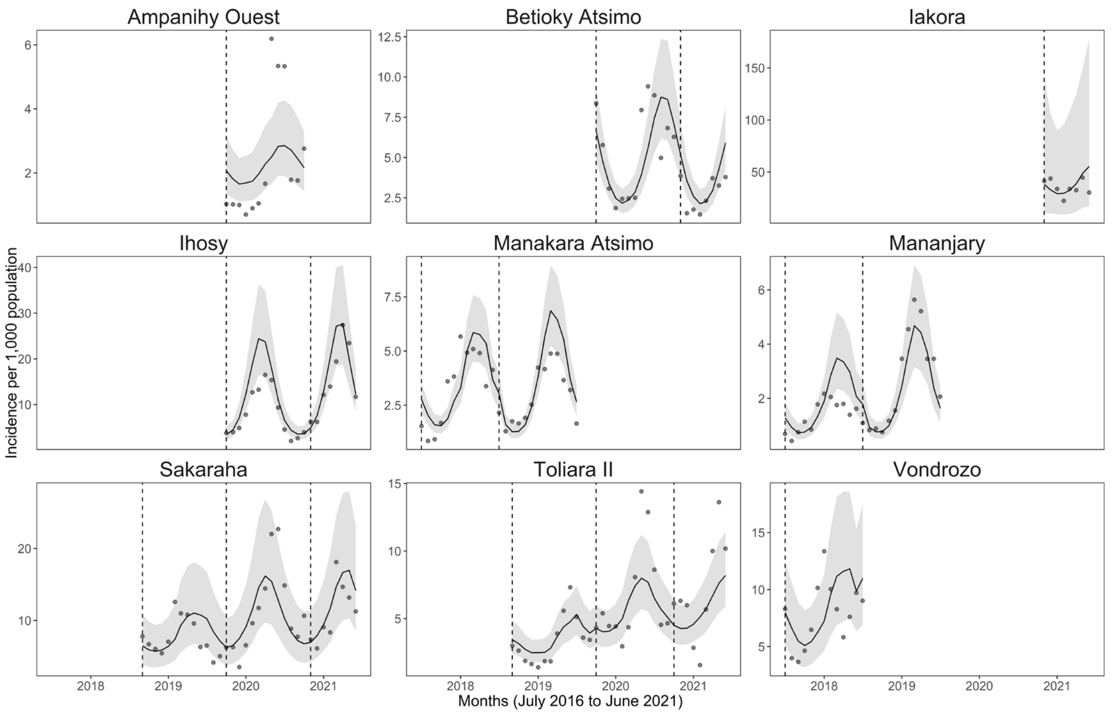

**B.**

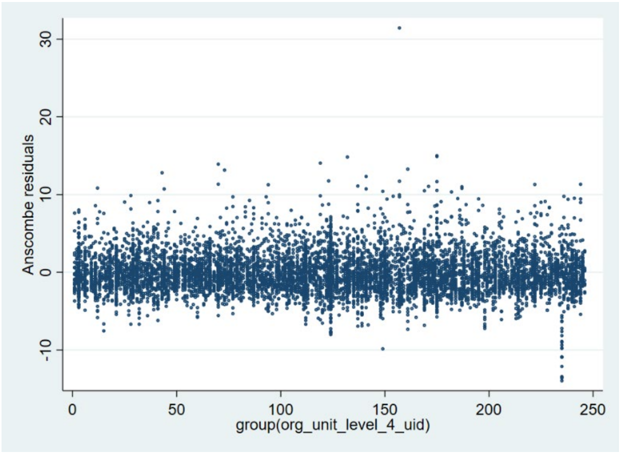
